# Comprehensive proteomics reveals stage-specific signatures in blood that precede diabetic kidney disease onset and progression

**DOI:** 10.64898/2026.11.10.26367534

**Authors:** Stefan Mutter, Emma Dahlström, Xiaoqi Luo, Joanna M. Kouki, Wenjun Ju, Viji Nair, Erkka Valo, Anna Syreeni, Zeinab Abdelrahman, Claire Hill, Laura Smyth, Heather R. Jackson, Andrew Parton, Valma Harjutsalo, Katalin Susztak, Alexander P. Maxwell, Amy Jayne McKnight, Gareth McKay, Matthias Kretzler, Per-Henrik Groop, Niina Sandholm, the FinnDiane Study Group

## Abstract

Diabetic kidney disease is the leading cause of kidney failure worldwide, yet the molecular events underlying disease initiation and progression remain unclear. We measured 5,420 circulating proteins in 830 adults with type 1 diabetes and examined their longitudinal associations with onset and progression of kidney disease over 10 years of follow-up. We identified 130 proteins associated with at least one outcome independent of established clinical risk factors, of which 59 replicated in an independent cohort. HAVCR1/KIM-1, GFRA1, CD14 and GDF15 predicted albuminuria onset up to a decade before detectable disease. Albuminuria progression pointed to the kidney–liver axis (ASGR1, RBP4), whereas rapid kidney function decline and incident kidney failure pointed to immune and tumor necrosis factor receptor signaling. Mendelian randomization prioritized UMOD, SORT1, CLSPN, MAST4 and EGF as potential targets. These findings reveal molecular perturbations preceding clinical manifestations of diabetic kidney disease and identify distinct pathways associated with disease progression.

---

Diabetic kidney disease (DKD) is the leading cause of chronic kidney disease (CKD) and kidney failure (KF) worldwide, affecting a considerable proportion of the 537 million adults living with diabetes^1,2^. Despite substantial advances, the clinical course of DKD is heterogeneous: some individuals maintain stable kidney function for decades whereas other progress rapidly to KF despite similar clinical risk factors^3^. Understanding this variability and identifying early markers for disease onset and progression is therefore needed.

Accumulating evidence suggests that DKD develops years before onset of clinically apparent disease. Alterations in lipid metabolism, central obesity and insulin resistance are detectable long before albuminuria develops in individuals with diabetes^4^. These systemic changes are likely accompanied by alterations in the circulating proteome, offering early insights into disease pathophysiology and risk. However, most existing proteomic studies of DKD have focused on its most advanced stages^5,6^, identifying markers such as kidney injury molecule-1 (KIM-1, also known as HAVRC1)^7^ and several members of TNF receptor family that predict progression to KF^5,8^. Comparatively little is known about the proteomic signature of albuminuria onset, or the earlier transition from moderate to severe albuminuria. This lack of early markers is a critical gap, since clinical trials have consistently demonstrated that early intervention yields substantially greater long-term benefits than treatment initiated after severe kidney damage is established ^9,10,11^.

Defining these early molecular changes is particularly important in type 1 diabetes where diabetes onset typically occurs early in life and complications therefore disproportionately contribute to loss of healthy life-years. Despite declining incidence rates of severe DKD in type 1 diabetes, a considerable burden of kidney disease persists^12^.

Recent advances in high-throughput proteomics enable comprehensive and unbiased profiling of the circulating proteome, providing new opportunities to systematically identify biomarkers of DKD risk at all stages of disease to uncover biological pathways.

In one of the most comprehensive proteomic studies of DKD to date, we quantified 5,420 circulating proteins in 830 adults with type 1 diabetes from the Finnish Diabetic Nephropathy (FinnDiane) Study (**Fig. 1**). Revealing widespread cross-sectional associations between kidneys and the circulating proteome, importantly, we also identified proteins predicting albuminuria onset, albuminuria progression, progression to KF and rapid kidney function decline. Lead proteins were independently validated in 249 additional participants with type 1 or type 2 diabetes. Calculating protein quantitative trait loci (pQTLs) and integrating these genome-wide findings with Mendelian randomization (MR) provided for causal evidence. Altogether, we identified both established and previously unrecognized markers, including several proteins detectable before clinical onset of albuminuria. They further implicate inflammation and the kidney–liver axis connecting DKD stages that have previously been studied in isolation.

**Figure 1.**
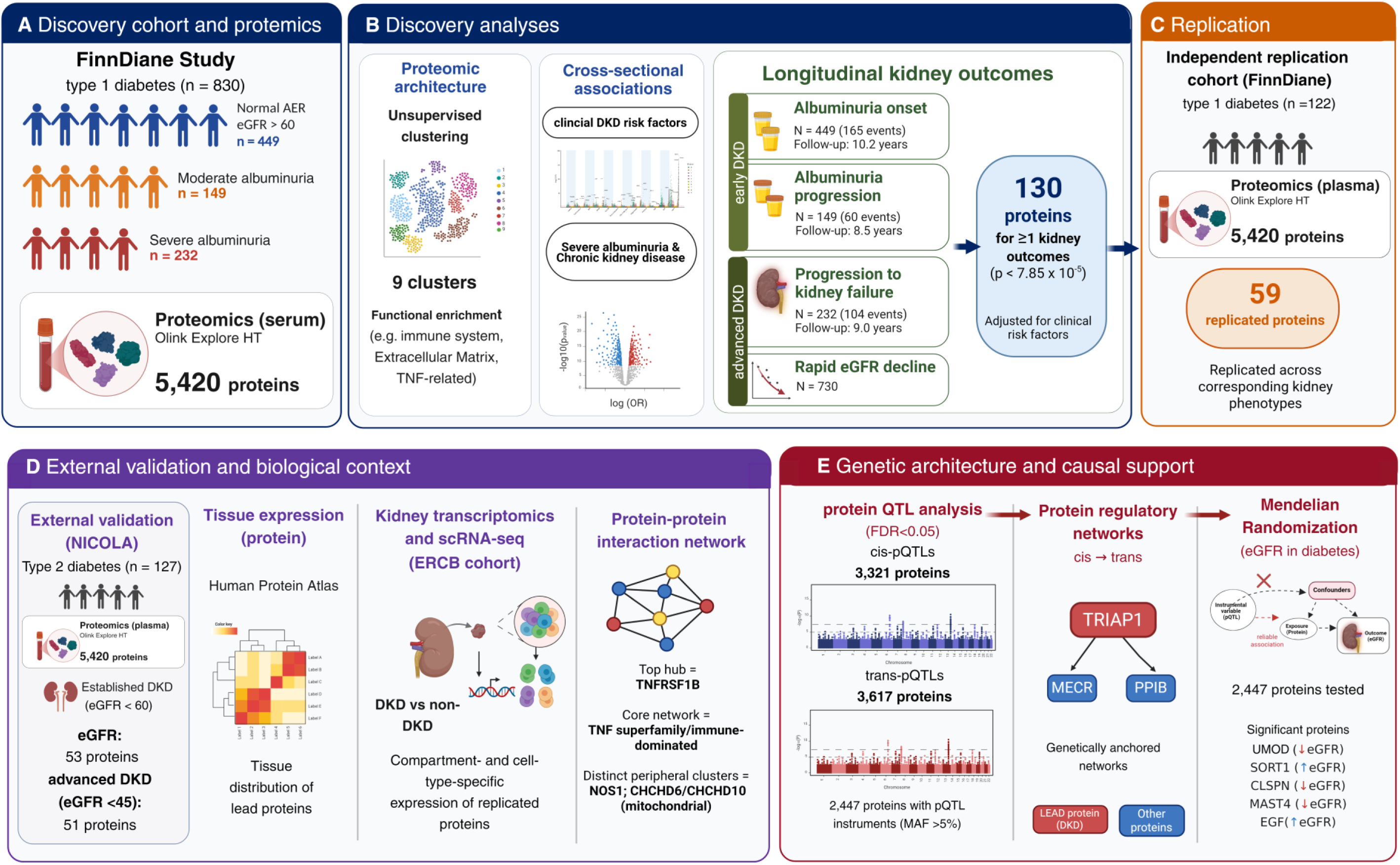
Study Design and analytical workflow. Proteomic profiling was performed in 830 adults with type 1 diabetes from the FinnDiane Study using the Olink Explore HT platform, quantifying 5,420 circulating proteins. Unsupervised clustering identified distinct proteomic architectures, and cross-sectional analyses evaluated associations with clinical risk factors for DKD and prevalent severe albuminuria and chronic kidney disease. Longitudinal analyses examined associations between circulating proteins and incident albuminuria, albuminuria progression, kidney failure, and rapid decline in estimated glomerular filtration rate (eGFR). A total of 130 proteins were associated with one or more kidney outcomes after adjustment for clinical risk factors. Findings were evaluated in an independent FinnDiane replication cohort (n=122), where 59 proteins replicated across corresponding kidney phenotypes. Biological validation incorporated external clinical, tissue-level, and single-cell transcriptomic datasets, including the Human Protein Atlas and the ERCB kidney transcriptomics cohort. Genetic analyses mapped cis- and trans-protein quantitative trait loci (pQTLs), reconstructed protein regulatory networks, and applied Mendelian randomization to investigate putative causal effects of circulating proteins on DKD. FinnDiane: Finnish Diabetic Nephropathy Study, GWAS: genome-wide association analysis, pQTL: protein quantitative trait locus, CKD: chronic kidney disease. Follow-up times shown are mean years.

## RESULTS

### Study population and proteomic landscape

The discovery cohort included 830 adults with type 1 diabetes from the FinnDiane Study **(Supplementary Fig. 1**): 449 individuals had a normal albumin excretion rate (AER) and estimated glomerular filtration rate (eGFR) above 60 mL/min/1.73m^2^, while 149 had moderate and 232 severe albuminuria. Compared to those with a normal AER, individuals with albuminuria were more frequently obese, had higher systolic blood pressure and more chronic inflammation (**Table 1**). The Olink Explore HT assay covered 5,420 circulating proteins. Unsupervised clustering identified nine protein clusters with partially distinct functional enrichments (**Extended Data Fig. 1**). The largest cluster (cluster 7, n=1,467 proteins) did not represent any significant enrichment, whereas the other clusters were more specific, ranging from immune system and lipid metabolism (cluster 1,2, and 9), chromatin organization (cluster 6), to cellular stress responses and cell cycle regulation (cluster 4, 5).

**Table 1.** Baseline characteristics according to kidney health.

|  | Normal AER and CKD stage 1 or 2 | Moderate or severe albuminuria | P |
|---|---|---|---|
| Women (%) | 40.76 | 37.27 | 0.34 |
| Age (years) | 38.61 ± 11.60 | 43.31 ± 11.85 | 1.27 × 10<sup>-8</sup> |
| Diabetes duration (years) | 22.38 ± 11.27 | 30.62 ± 10.87 | 4.04 × 10<sup>-25</sup> |
| Age of diabetes onset (years) | 16.23 ± 9.94 | 12.69 ± 8.39 | 3.42 × 10<sup>-8</sup> |
| HbA1c (%) | 8.37 ± 1.50 | 8.57 ± 1.42 | 0.05 |
| BMI (kg/m<sup>2</sup>) | 25.39 ± 3.64 | 26.64 ± 4.39 | 1.26 × 10<sup>-5</sup> |
| Waist-height ratio | 0.50 ± 0.06 | 0.54 ± 0.07 | 3.90 × 10<sup>-15</sup> |
| Total cholesterol (mmol/L) | 4.78 ± 0.83 | 4.90 ± 1.03 | 0.07 |
| HDL cholesterol (mmol/L) | 1.44 ± 0.39 | 1.38 ± 0.43 | 0.05 |
| LDL cholesterol (mmol/L) | 2.87 ± 0.79 | 2.87 ± 0.91 | 0.99 |
| Triglycerides (mmol/L) | 0.94 (0.73, 1.32) | 1.30 (0.93, 1.91) | 2.99 × 10<sup>-17</sup> |
| hs-CRP<sup>†</sup> (mg/L) | 2.02 ± 1.79 | 2.42 ± 2.26 | 7.20 × 10<sup>-3</sup> |
| Systolic blood pressure (mmHg) | 133 ± 17 | 142 ± 19 | 1.72 × 10<sup>-12</sup> |
| Diastolic blood pressure(mmHg) | 79 ± 9 | 80 ± 11 | 0.05 |
| Smoking (%) | 26.98 | 29.21 | 0.54 |
| Antihypertensive medication (%) | 21.57 | 85.98 | 3.28 × 10<sup>-75</sup> |
| Statins (%) | 14.35 | 41.80 | 1.77 × 10<sup>-18</sup> |
| eGFR (mL/min/1.73m<sup>2</sup>) | 107 ± 17 | 77 ± 34 | 4.10 × 10<sup>-47</sup> |
| Moderate albuminuria (%) | 0.00 | 39.11 | 6.92 × 10<sup>-48</sup> |
| Severe albuminuria (%) | 0.00 | 60.89 | 7.34 × 10<sup>-84</sup> |

Normally distributed data are presented as mean ± standard deviation, otherwise median (IQR). Counts as percentages. <sup>†</sup>Individuals with acute inflammation defined by hs-CRP ⩾ 10 mg/L excluded. CKD: chronic kidney disease.

### Cross-sectional protein associations with cardio-kidney-metabolic traits

Circulating protein associations with baseline clinical traits were dominated by AER and eGFR which showed the largest number of significant associations and the strongest statistical evidence (**Extended Data Fig. 2, Supplementary Table 1** and https://finndiane.shinyapps.io/Protein_Association/). Proteins involved in lipid metabolism, including LDLR were strongly associated with triglycerides and FABP4 with measures of obesity. In addition, inflammatory makers such as LRG1 and FGL1 were associated with hs-CRP (high-sensitivity C-reactive protein).

In binary analyses, consistent with above continuous measures of kidney health, severe albuminuria and CKD (eGFR<60 mL/min/1.73m²) were associated with 28% (n=1,535) and 29% (n=1,576) of all tested proteins, respectively (P<7.85×10^-5^; **Supplementary Table 2**). Of these, 9% (n=484) remained significant after adjustment for eGFR in severe albuminuria, and 18% (n=959) after adjustment for severe albuminuria in CKD (**Fig. 2a,b**). Of note, 49% of individuals with severe albuminuria had a preserved eGFR, whereas 92% of those with CKD had severe albuminuria. Protein associations between both group were moderately correlated (Spearman ρ=0.37, P<2.2×10^-16^), suggesting partially distinct signatures. Eight Reactome pathways were enriched in both, including *extracellular matrix organization*, *TNF receptor binding* and *neutrophil degranulation* (**Fig. 2c, Supplementary Table 3**).

**Figure 2.**
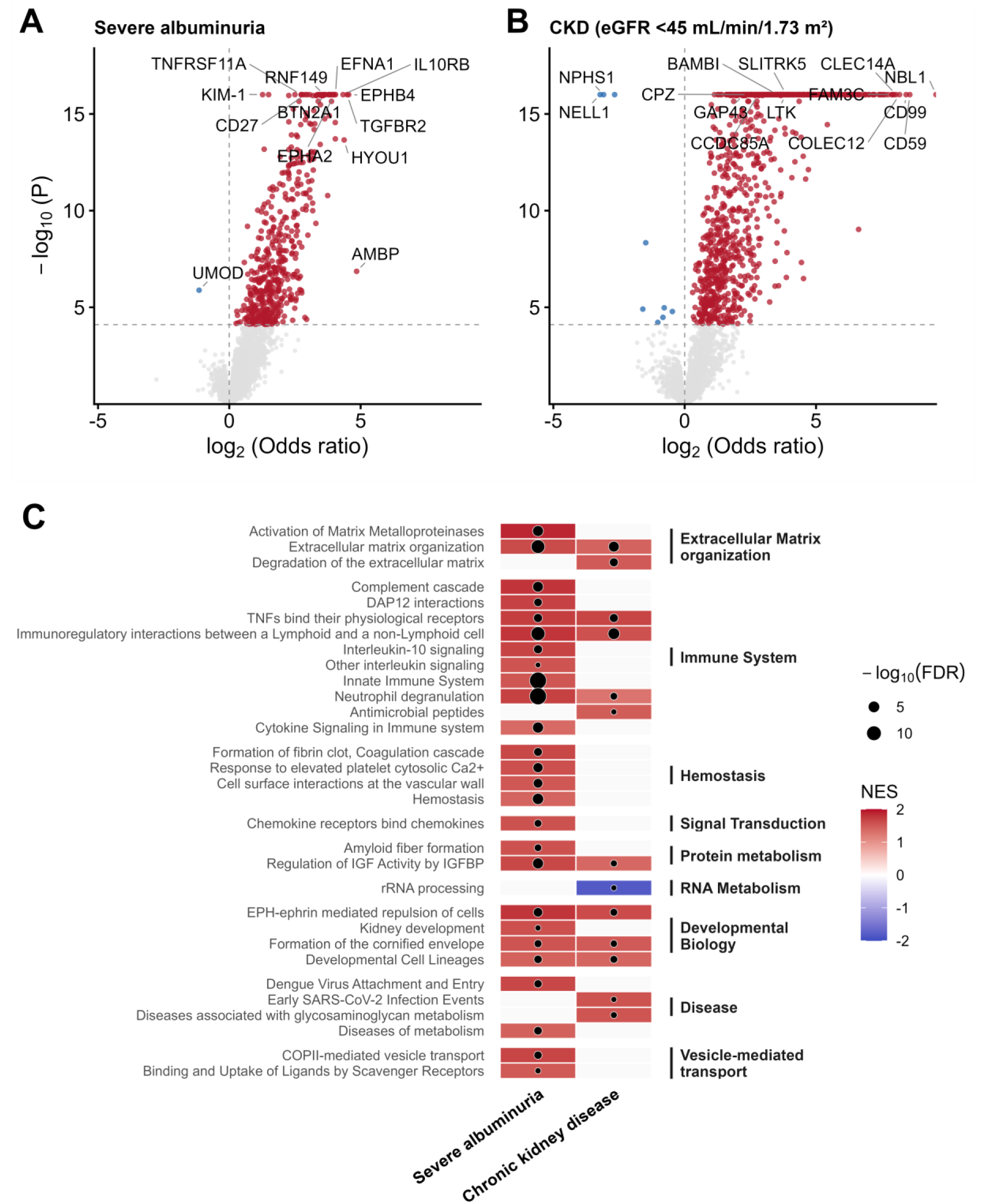
Cross-sectional associations with baseline diabetic kidney disease. (A) Odds ratios for severe albuminuria vs. moderate albuminuria or normal AER adjusted for sex, eGFR and technical covariates. (B) Odds ratios for chronic kidney disease (CKD), adjusted for sex, severe albuminuria and technical covariates. (C) Reactome pathways (FDR<0.05) from gene set enrichment analysis (E) of proteins ranked according to their associations with severe albuminuria or CKD in minimally adjusted models. Color represents the normalized enrichment score (NES), normalized to account for differences in pathway size. Positive values (red) indicate enrichment among proteins whose higher abundance was associated with increased risk, negative values (blue) indicate enrichment among proteins whose higher abundance was associated with decreased risk. Point size indicate pathway significance (-log10(FDR)).

### Survival analysis identifies 130 proteins associated with albuminuria onset, its progression, progression to KF and rapid kidney function decline

We next conducted competing risk survival analysis accounting for death to identify proteins associated with albuminuria onset (N=449, 165 events, follow-up: 10.2 ± 5.4 years), progression of albuminuria (N=149, 60 events, 8.5 ±5.0 years), and progression to KF (N=232, 104 events, 9.0 ± 5.2 years; **Supplementary Table 4**). In addition, we used Firth’s bias-reduced logistic regression to identify proteins associated with rapid kidney function decline (annual eGFR loss >3mL/min/1.73m^2^, N=730, 145 cases).

In minimally adjusted models, 38 proteins were proteome-wide significantly (P<7.85×10^-5^) associated with albuminuria onset, 50 with albuminuria progression, 955 with progression to KF and 205 with rapid kidney function decline (**Supplementary Table 5**). Functional enrichment analysis revealed four pathways enriched in all outcomes (false discover rate (FDR) <0.05) with similar enrichment scores, namely *extracellular matrix organization, innate immune system, immunoregulatory interactions and regulations of insulin-like growth factor* (**Fig. 3e**, **Supplementary Table 3**). Onset and albuminuria progression shared three further pathways (*diseases of metabolism, neutrophil degranulation,* and *plasma lipoprotein assembly remodeling and clearance*), while albuminuria progression, progression to KF and rapid decline shared the pathway *cell surface interactions at the vascular wall*. The biggest overlap was found in pathways for progression to KF and rapid decline with altogether 11 pathways including *TNF receptor binding, cytokine signaling in the immune system* and *interleukin 10 signaling*.

**Figure 3.**
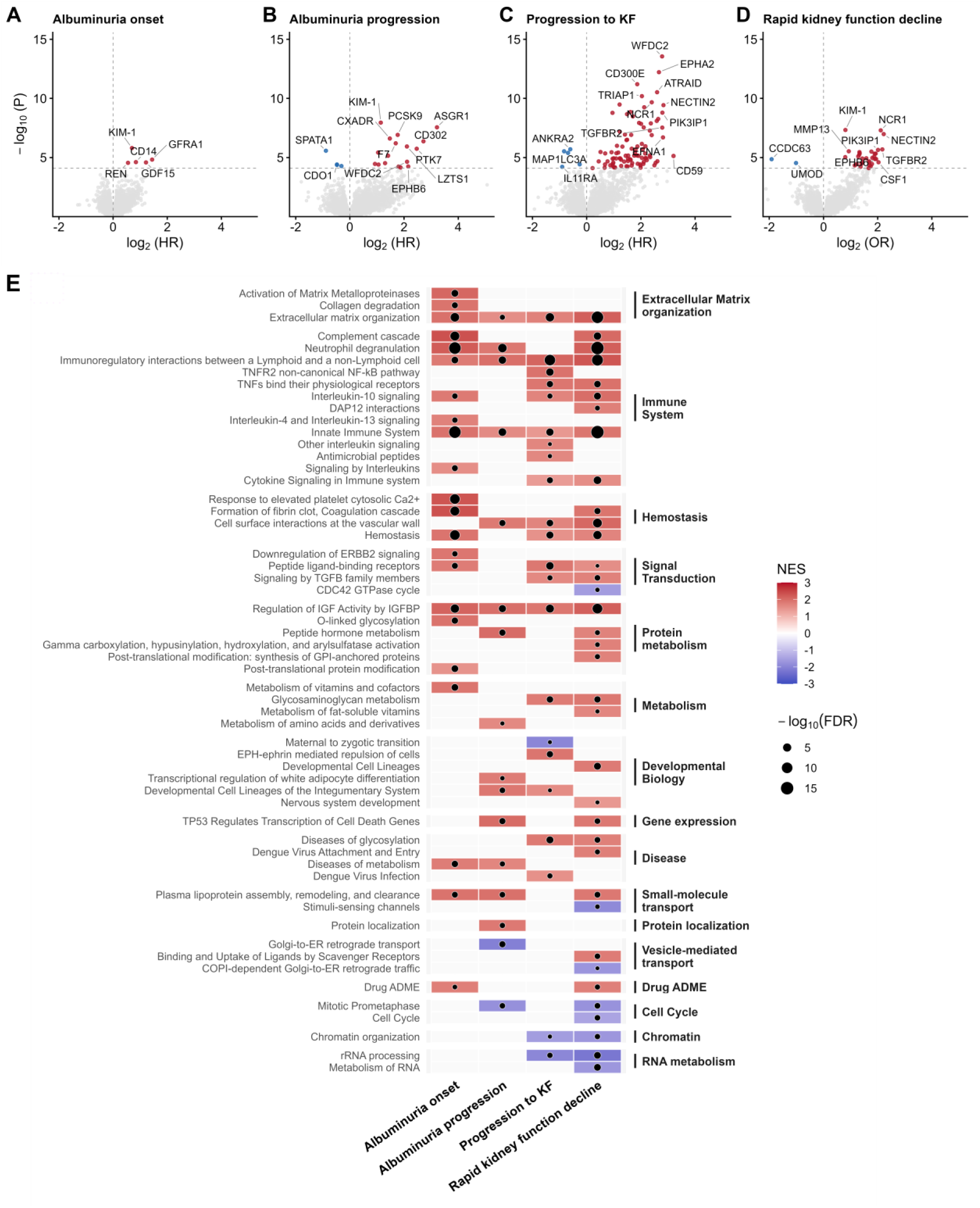
Proteins predicting kidney disease onset and progression. Subdistribution hazard ratios (sHR) and P values for onset of albuminuria (A), albuminuria progression (B), and progression to kidney failure (C); and odd ratios for rapid kidney function decline (D) in models adjusted for the clinical risk factors. Red and blue colors indicate proteins proteome-wide significantly associated (P<7.85×10^-5^) with the outcome, also indicated with the dashed horizontal line. Reactome pathways (FDR<0.05) from gene set enrichment analysis (E) of proteins ranked according to their associations with each kidney outcomes in minimally adjusted models. Color represents the normalized enrichment score (NES), normalized to account for differences in pathway size. Positive values (red) indicate enrichment among proteins whose higher abundance was associated with increased risk, negative values (blue) indicate enrichment among proteins whose higher abundance was associated with decreased risk. Point size indicates pathway significance (-log10(FDR)).

Representing putative biomarkers independent of known clinical risk factors, we identified 130 proteins that were proteome-wide significantly associated with at least one of the four longitudinal outcomes after adjusting for sex, age, age at diabetes onset, eGFR, HbA1c, triglycerides, systolic blood pressure, waist-height ratio, hs-CRP and current smoking status. We further evaluated these proteins in an independent replication cohort of 122 additional participants with type 1 diabetes with their plasma proteome measured using the same platform. Discovery cohort outcomes were matched to their closest available one: albuminuria onset to baseline albuminuria, albuminuria progression to severe baseline albuminuria, rapid function decline to baseline CKD stages 3–4, and KF progression to a composite, longitudinal endpoint of incident KF or ≥40% eGFR decline. Overall, 59 out of 130 proteins showed evidence of replication (**Supplementary Table 6**).

Regarding albuminuria onset, five proteins (KIM-1, GFRA1, CD14, GDF15, REN) were proteome-wide significant after adjusting for clinical risk factors (**Fig. 3a**). KIM-1, GFRA1, CD14 and GDF15 replicated (P<0.01, **Table 2**). Among them, KIM-1 was consistently associated with all four kidney outcomes. Of note, CD14 and GDF15 were specifically associated with albuminuria onset and not with the later stages of DKD progression, whereas GFRA1 was also associated with rapid decline. Investigation of the associated cardiometabolic intermediate traits showed that serum CD14 abundance correlated most strongly with hs-CRP, GFRA1 with type 1 diabetes duration; and GDF15 with age (**Extended Data Fig. 3**).

**Table 2.** Significant proteins (P<7.85 × 10^-5^) predicting onset of albuminuria and albuminuria progression in models adjusted for clinical risk factors.

|  |  |  | Discovery |  | Replication |  |
|---|---|---|---|---|---|---|
| Protein | UniProt ID | Outcome | sHR [95% CI] | P | OR [95% CI] | P |
| KIM-1 | Q96D42 | Alb. Onset | 1.62 [1.33, 1.97] | 1.60×10<sup>-6</sup> | 1.58 [1.15, 2.26] | 3.34×10<sup>-3</sup>† |
| GFRA1 | P56159 | Alb. Onset | 2.72 [1.73, 4.27] | 1.49×10<sup>-5</sup> | 2.01 [1.21, 3.47] | 6.50×10<sup>-3</sup>† |
| CD14 | P08571 | Alb. Onset | 1.79 [1.37, 2.35] | 2.43×10<sup>-5</sup> | 2.30 [1.33, 4.23] | 2.27×10<sup>-3</sup>† |
| GDF15 | Q99988 | Alb. Onset | 2.32 [1.57, 3.44] | 2.59×10<sup>-5</sup> | 3.57 [1.95, 7.16] | 6.62×10<sup>-6</sup>† |
| REN | P00797 | Alb. Onset | 1.47 [1.23, 1.75] | 2.80×10<sup>-5</sup> | 1.20 [0.94, 1.56] | 0.15 |
| KIM-1 | Q96D42 | Alb. progression | 2.21 [1.68, 2.89] | 1.13×10<sup>-8</sup> | 1.46 [1.13, 1.94] | 3.46×10<sup>-3</sup> |
| ASGR1 | P07306 | Alb. progression | 9.27 [4.22, 20.34] | 2.81×10<sup>-8</sup> | 2.39 [1.41, 4.34] | 6.92×10<sup>-4</sup>† |
| PCSK9 | Q8NBP7 | Alb. progression | 3.40 [2.16, 5.36] | 1.23×10<sup>-7</sup> | 1.42 [0.71, 2.93] | 0.32 |
| CXADR | P78310 | Alb. progression | 2.78 [1.89, 4.10] | 2.50×10<sup>-7</sup> | 2.76 [1.70, 4.75] | 1.60×10<sup>-5</sup>† |
| CD302 | Q8IX05 | Alb. progression | 6.58 [3.17, 13.66] | 4.32×10<sup>-7</sup> | 2.33 [1.42, 4.04] | 6.21×10<sup>-4</sup>† |
| F7 | P08709 | Alb. progression | 3.23 [2.04, 5.13] | 6.36×10<sup>-7</sup> | 1.46 [0.74, 2.99] | 0.28 |
| LZTS1 | Q9Y250 | Alb. progression | 4.31 [2.39, 7.77] | 1.18×10<sup>-6</sup> | 2.99 [1.81, 5.43] | 3.87×10<sup>-6</sup>† |
| PTK7 | Q13308 | Alb. progression | 5.54 [2.74, 11.19] | 1.84×10<sup>-6</sup> | 4.84 [2.29, 11.64] | 7.15×10<sup>-6</sup>† |
| SPATA1 | Q5VX52 | Alb. progression | 0.54 [0.42, 0.70] | 2.71×10<sup>-6</sup> | 0.92 [0.73, 1.16] | 0.49 |
| RBP4 | P02753 | Alb. progression | 2.06 [1.52, 2.80] | 3.83×10<sup>-6</sup> | 2.05 [1.34, 3.29] | 6.85×10<sup>-4</sup>† |
| DKKL1 | Q9UK85 | Alb. progression | 2.66 [1.74, 4.07] | 7.06×10<sup>-6</sup> | 1.95 [1.12, 3.57] | 0.02 |
| WFDC2 | Q14508 | Alb. progression | 4.32 [2.20, 8.51] | 2.27×10<sup>-5</sup> | 3.16 [1.88, 5.67] | 3.83×10<sup>-6</sup>† |
| MMP7 | P09237 | Alb. progression | 2.46 [1.61, 3.76] | 2.98×10<sup>-5</sup> | 2.22 [1.41, 3.67] | 3.54×10<sup>-4</sup>† |
| NDST1 | P52848 | Alb. progression | 1.90 [1.40, 2.58] | 3.46×10<sup>-5</sup> | 1.12 [0.79, 1.61] | 0.53 |
| CDO1 | Q16878 | Alb. progression | 0.72 [0.61, 0.84] | 3.86×10<sup>-5</sup> | 0.54 [0.37, 0.75] | 8.88×10<sup>-5</sup>† |
| PRSS2 | P07478 | Alb. progression | 2.08 [1.47, 2.95] | 3.92×10<sup>-5</sup> | 2.13 [1.38, 3.43] | 4.11×10<sup>-4</sup>† |
| CENPS | Q8N2Z9 | Alb. progression | 0.72 [0.61, 0.84] | 4.24×10<sup>-5</sup> | 0.94 [0.75, 1.15] | 0.53 |
| KIF13B | Q9NQT8 | Alb. progression | 0.81 [0.73, 0.89] | 5.20×10<sup>-5</sup> | 0.78 [0.55, 1.09] | 0.15 |
| EPHB6 | O15197 | Alb. progression | 4.46 [2.15, 9.23] | 5.71×10<sup>-5</sup> | 4.39 [2.35, 9.23] | 3.80×10<sup>-7</sup>† |
| PRSS8 | Q16651 | Alb. progression | 3.48 [1.89, 6.39] | 5.83×10<sup>-5</sup> | 1.79 [1.10, 3.00] | 0.02 |
| PROC | P04070 | Alb. progression | 3.66 [1.93, 6.94] | 6.91×10<sup>-5</sup> | 1.25 [0.64, 2.49] | 0.51 |

UniProt ID: UniProt protein identifier. Alb.: Albuminuria. sHR [95% CI]: sub-Hazard Ratio and the 95% confidence interval for the outcome, accounting for death as the competing risk. Replication OR and P: Odds ratio and *P* from minimally adjusted Firth's bias-reduced logistic regression models in cross-sectional analysis of 122 independent adults with type 1 diabetes. †significant after correction for number of tested proteins per outcome (i.e. 5 for albuminuria onset; 21 for albuminuria progression)

Twenty-one proteins were proteome-wide significantly associated with the progression of albuminuria after adjustment for the clinical risk factors, of which 11 were replicated (**Table 2**, **Fig. 3b**). KIM-1 was the most significant predictor (P=1.13×10^-8^) and the protein with the largest effect size was ASGR1 (sHR=9.27, P=2.81×10^-8^). Stratification by ASGR1 abundance produced clearly separated cumulative incidence curves (**Supplementary Fig. 2**). While correlated with triglycerides (**Extended Data Fig. 3**), it showed predominant expression in the liver as opposed to the kidneys in tissue expression data from the Human Protein Atlas (**Extended Data Fig. 4b**). Similarly, RBP4 was associated with triglycerides and expressed in the liver, but also in the kidneys (**Extended Data Fig. 4b**) with additional evidence of overexpression in CKD especially in injured proximal tubule cells (P=1.3×10^-31^) in kidney single-cell RNA sequencing (scRNAseq) data^13^ (**Supplementary Table 7**). In general, hierarchical clustering in the Human Protein Atlas clustered kidney and liver together, consistent with a potential involvement of the kidney–liver axis in albuminuria progression.

Of the 98 proteins proteome-wide significantly associated with progression to KF in the clinically adjusted model (**Fig. 3c, Supplementary Table 6**), 30 were replicated (P<5.1×10^-4^; **Fig. 4**). Furthermore, of the 34 proteins associated with rapid kidney function decline (**Fig. 3d**), 30 replicated (P<0.0015; **Supplementary Table 6, Fig. 4**). Of note, 59% of the proteins associated with rapid decline were also associated with progression to KF. Significant proteins for both outcomes showed a strong correlation with baseline eGFR, except for MMP13 which correlated with sex (**Extended Data Fig. 3**).

**Figure 4.**
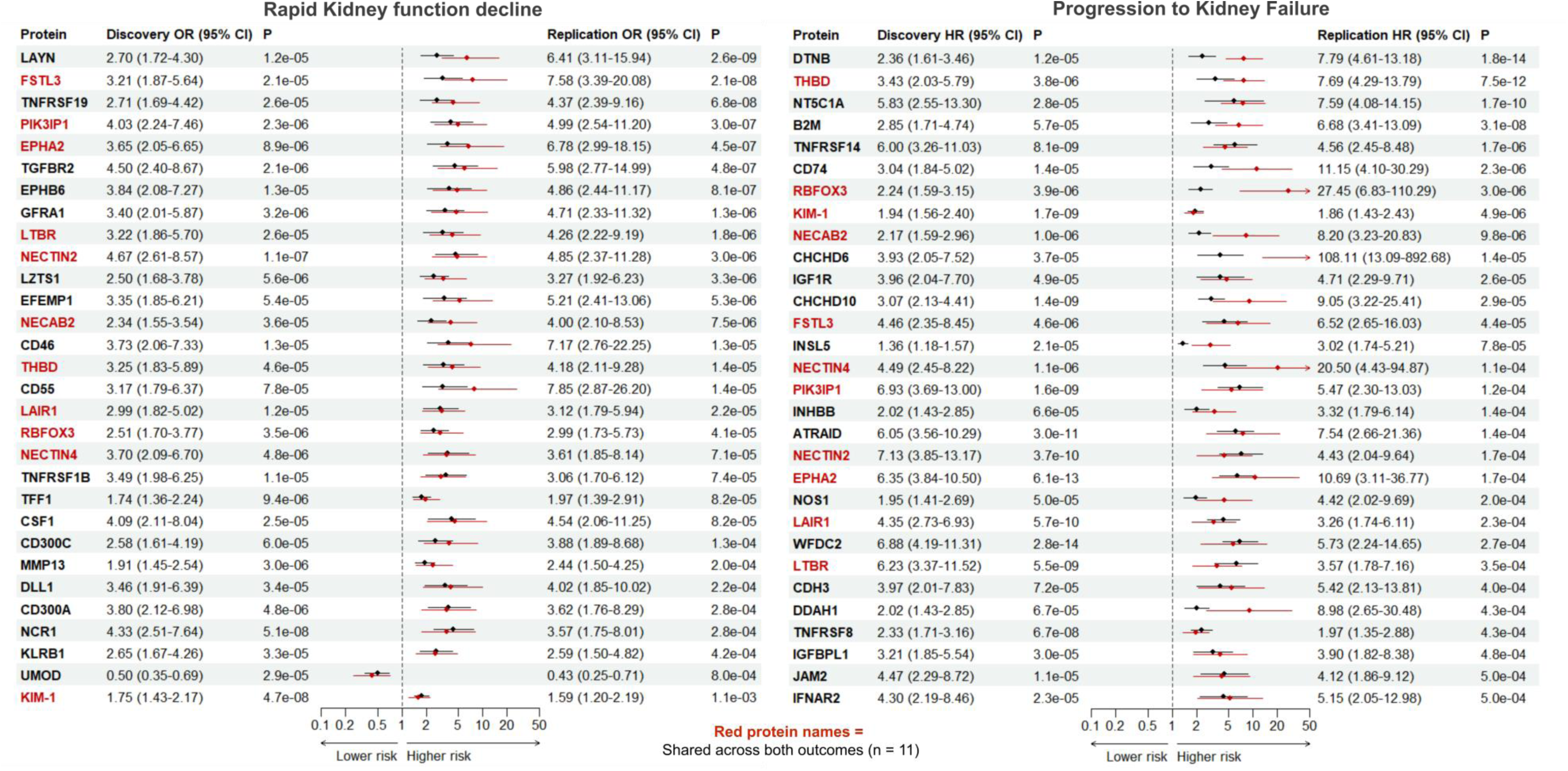
Forrest plot of replicated proteins for progression to kidney failure and rapid kidney function decline. Forest plots show effect estimates and 95 % confidence intervals (CI) for proteins identified in the discovery analysis and subsequently replicated in an independent cohort (n=122, rapid eGFR decline: P < 0.0015; kidney failure: P < 5.1 × 10⁻⁴). Black circles indicate discovery estimates and red indicate the replication estimates. Proteins are ordered by replication P value.

To assess whether replicated associations were robust to sample storage conditions, we evaluated protein stability in a subset of 20 individuals with parallel serum samples stored at −20 °C and −80 °C. The 59 replicated proteins showed high correlations across storage conditions (median Pearson ρ=0.94 [interquartile range 0.91-0.96]; **Supplementary Table 8**).

### Independent validation of replicated proteins in kidney disease in type 2 diabetes

To assess the generalizability of the findings, we tested whether the replicated 59 proteins were also associated with eGFR or advanced DKD (eGFR < 45mL/min/1.73m^2^) in 127 NICOLA study participants with type 2 diabetes and eGFR <60mL/min/1.73m^2^. Of the 59 replicated proteins, 53 (90%) were associated with eGFR and 51 (86%) with advanced DKD, while 49 (83%) were associated with both outcomes (FDR<0.05; **Supplementary Table 9).** Effect directions were highly consistent across diabetes types, demonstrating substantial overlap between diabetes types.

### Kidney expression evidence

Based on kidney biopsies from individuals with DKD (N=17 and 12 for tubular and glomerular, respectively) and healthy living donors (N=30 and 31 for tubular and glomerular, respectively), we investigated altered kidney gene expression in the European Renal cDNA Bank (ERCB)^14–16^. Out of 15 genes, 6 (40%) were significantly differentially regulated (P<0.05/15) in either glomerular or tubulointerstitial compartment and four of them (CD14, CXADR, MMP7, WFDC2) in both compartments. (**Supplementary Table 10**). This suggests that the expression and regulation of these genes in the kidney are broadly consistent with those of their corresponding proteins in circulation.

To find further evidence (FDR<0.05) for kidney expression for the 59 replicated proteins, we queried scRNAseq data in in healthy controls and individuals with CKD from the Human Kidney Single Cell Transcriptome dataset^13^ (*Susztak & KPMP merged*, https://susztaklab.com/hk_genemap_kpmp/scRNA, **Supplementary Table 7**) where 53 out of the 59 proteins showed evidence of cell type specific expression in the kidneys including immune cells. Furthermore, 38 of those were additionally differentially expressed in CKD including earlier stage markers such as CD14, GDF15, CXADR, RBP4 and later stage markers such as MMP7, WFDC2, PIK3IP1 and UMOD.

### Interaction network of replicated proteins associated with deteriorating kidney health

Protein-protein interaction (STRING database) networks of the 59 replicated proteins (**Extended Data Fig. 5, Supplementary Table 11**) indicated TNFRSF1B as the most central protein in the network, exhibiting the highest degree (degree of 8), betweenness and eigenvector centrality. TNFRSF14 and BM2 had a degree of 7, followed by GDF15 and TNFRSF8 (degree of 6), and NECTIN2 (degree of 5). Overall, the largest connected component was dominated by immune-related proteins, particularly the members of the TNF superfamily, whereas two smaller separate networks consisted of NOS1 in its center and one with two mitochondrial proteins CHCHD6 and CHCHD10.

### Genetic architecture of proteins and kidney disease-associated regulatory networks

To characterize the genetic regulation of circulating protein levels measured on the Olink Explore HT platform, we performed genome-wide analyses of both local (*cis*-pQTL; within 1 Mb) and distant (*trans*-pQTLs) genetic effects. We identified altogether 325,772 *cis*-pQTLs for 3,231 proteins (60.0%), and 97,709 trans-pQTLs for 3,617 proteins (67.0%), at FDR<0.05, demonstrating extensive genetic control of the circulating proteome (**Extended Data Fig. 6a, Supplementary Table 12**). Among proteins with significant pQTLs, nearly half (47%) showed evidence of both local and distal genetic regulation (**Extended Data Fig. 6b-d**). Several proteins showed particularly strong *cis* effects, with lead variants explaining more than 50% of protein level-variance (*r*^2^>0.5;). Notably, 1,300 proteins with *cis*-pQTLs, and 1,830 proteins with *trans*-pQTLs proteins were unique to the Olink Explore HT platform compared to earlier Olink panels, indicating that newly assayed proteins are also under strong genetic control^17^.

To explore potential regulatory mechanisms among the 130 DKD-associated proteins, we inferred interactions based on *cis*-and *trans*-pQTL associations (FDR<0.1, minor allele frequency (MAF)≥0.05). Specifically, when a *cis*-pQTL variant for the lead protein acts as a *trans*-pQTL variant for another protein, the *trans*-regulated protein was considered a potential downstream target of the *cis*-regulated protein. Notably, this approach identified endothelial protein C receptor (PROCR) as an upstream regulator of the albuminuria progression-associated PROC through the shared pQTL variant rs6579210. This variant was also reported as a *cis*-eQTL for PROCR in eQTLGen^18^. Furthermore, among others, we identified TRIAP1, associated with progression to KF, as a putative upstream regulator of PPIB and MECR (**Extended Data Fig. 7**).

### Mendelian randomization

To evaluate genetic support for potential causal effects of circulating protein abundances, we performed MR analyses using identified pQTLs as instruments for the proteins and summary statistics from a genome-wide association study (GWAS) meta-analysis on eGFR in diabetes as outcome^19^.

Among 2,447 proteins with available pQTLs (MAF≥5%), higher genetically predicted levels of UMOD were associated with lower eGFR, and higher SORT1 levels with higher eGFR (FDR<0.05**, Supplementary Table 13).** In analyses restricted to individuals with diabetes but without severe albuminuria, CLSPN, MAST4, EGF and SORT1 were additionally associated with eGFR (FDR<0.05). Comparison with observational findings demonstrated both concordant (EGF, SORT1) and discordant effects as UMOD demonstrated opposing effect directions (**Supplementary Table 13)**. In contrast, CLSPN and MAST4 exhibited significant MR associations despite limited observational evidence.

Within the 130 lead proteins, 76 had suitable pQTL instruments for causal evaluation (**Supplementary Table 14**). In addition to significant MR evidence for UMOD, nominally significant (P<0.05) and directionally consistent evidence was observed for five of the 59 replicated proteins, including CD300A, ATRAID and INHBB for progression to KF and CD300A, CD55 and UMOD for rapid kidney function decline. Further nominal MR evidence was found for ADAMTSL2 and SPINK4.

## DISCUSSION

In this large-scale prospective study, covering 5,420 proteins measured in 830 adults with type 1 diabetes, we identified distinct molecular signatures across the continuum of DKD development and progression, including 130 proteins associated with onset of albuminuria, albuminuria progression, progression to KF, or rapid kidney function decline independent of well-established clinical risk markers. In an independent cohort of 122 adults with type 1 diabetes, 59 were replicated and 55 of those associated with advanced DKD or eGFR in 127 individuals with type 2 diabetes showing substantial transferability across diabetes types. By integrating proteomics data with pQTLs through MR, we provide some genetically anchored evidence of causality.

Notably, our study shows that proteomic changes are detectable well before albuminuria and eGFR become informative, when intervention could still alter the disease trajectory. Beyond KIM-1, a well-established marker^20–22^, we identified and replicated three proteins that predicted albuminuria onset independently of established risk factors: CD14, GFRA1, GDF15.

CD14, a co-receptor for bacterial lipopolysaccharide (LPS)^23^, supports a role for inflammatory processes at disease onset consistent with earlier findings of elevated circulating LPS in individuals with normal AER who subsequently developed albuminuria^24^. Soluble CD14 has been previously associated with DKD severity in type 2 diabetes^25^, and our study demonstrates its relevance at earlier stages. In our interaction network, CD14 was linked to CD55, a protein with potentially causal evidence (P<0.05) for rapid eGFR decline. CD55 regulates innate immunity^26^, one of the four pathways shared across all four longitudinal outcomes.

GFRA1 and GDF15 which were linked in the interaction network showed both expression in injured proximal tubule cells (P<1.5×10^−66^), and GDF15 additionally in principal and connecting tubule cells (P<10^-310^) with significant underexpression in CKD (P≤5.5×10^-89^). However, importantly, GDF15 is a ubiquitously expressed stress-response cytokine^27^. Previously, it has been linked to incident and worsening albuminuria in type 2 diabetes^28^, and to kidney function decline among individuals with type 1 diabetes and severe albuminuria^29^. GFRA1, implicated in kidney development^30^, was associated with albuminuria onset and rapid eGFR decline. An association with declining eGFR was also found in youth-onset type type 2 diabetes31. Together, these findings suggest that some of the proteins previously linked to established kidney dysfunction can be detected before albuminuria onset and that early molecular alterations may reflect both renal and systemic processes, indicating that key pathways are activated long before clinical disease.

The kidney–liver axis emerged as a prominent feature of earlier-stage outcomes, particularly albuminuria progression. For both albuminuria onset and progression, proteins in the plasma lipoprotein- and metabolism-related pathways were enriched. At the protein level, asialoglycoprotein receptor 1 (ASGR1), which had the largest effect size for progression, is predominantly expressed in liver hepatocytes and strongly correlated with triglycerides, consistent with its role as a putative drug target for hypercholesterolemia^32^. Its correlation with hs-CRP may reflect its reported link to coronary artery disease through inflammation^33^. RBP4, retinol-binding protein 4, which transports retinol from the liver to peripheral tissues, is expressed in both liver and the kidneys including proximal tubule, descending thin loop of Henle and parietal epithelial cells (P≤4.9×10^-43^). Although RBP4 is lost to urine in proteinuria, elevated circulating RBP4 has been observed in CKD linked to altered kidney–liver axis signaling^34^. This observation was supported in scRNAseq data as RBP4 was overexpressed in injured proximal tubule cells (P=1.3×10^-31^). RBP4 contributes to obesity-related inflammation, inhibits insulin signaling, promotes muscle insulin resistance, and its hepatic expression is linked to increased glucose production^35^. Although independent replication failed, PCSK9, predominantly liver-produced^36^, was among the lead signals for progression. Together, these findings point to a coordinated kidney–liver axis. Given the causal link between obesity and DKD^37^ and rising rates of obesity in type 1 diabetes^38^, the kidney–liver axis merits further exploration.

The albuminuria progression signals also link to kidney biology. Both WFDC2 and MMP7 were overexpressed in various tubular cells in CKD in the scRNAseq data, have been implicated in early renal fibrosis in type 2 diabetes and were collectively proposed as a kidney fibrosis panel^39^. Both markers were also associated with advanced DKD in our type 2 diabetes validation set. Furthermore, in mouse models, higher tubular expression of Mmp7 lead to glomerular injuries^40^. Soluble coxsackievirus and adenovirus receptor (CXADR) may promote albuminuria progression by inhibiting tight junction formation^41^. Consistent with this, Cxadr is expressed during podocyte injury in mice^42^. However, in the scRNAseq data, in CKD, CXADR was underexpressed in podocytes (P=2.2×10^-4^) and overexpressed in injured proximal tubule cells (P=9.3×10^-17^). Additionally, in our interaction network, it was directly linked with CD55 which had a potential causal role.

Progression to KF and rapid decline showed substantial overlap including immune and TNF-receptor signaling pathways. The shared lead signals converged with the 21-protein Joslin panel^8^ of which 14 proteins (CD160, CD27, DLL1, EFNA4, EPHA2, GFRA1, KIM-1, LAYN, LTBR, NECTIN4, PI3, TNFRSF1A, TNFRSF1B, WFDC2) were proteome-wide significant in our late-stage phenotypes. TNFRSF1B was the most central node in our interaction network, closely rivalled by onset marker GDF15, absent in the Joslin panel. NECTIN2, among the strongest later-stage associations, was central in a subnetwork that linked to NECTIN4, TNFRSF14, NCR1, CD55 and CD46, the latter two protecting podocytes from complement-mediated damage^43^. The urinary complement proteome has been associated with incident KF^44^. NECTIN2 itself was overexpressed in podocytes in individuals with CKD (P=1.2×10^-11^). Its expression during injury-repair states of the thick ascending limb of Henle’s loop^45^, together with its presence in the leading edge of the immunoregulation pathway shared across all four longitudinal outcomes further support immune system involvement.

Moreover, we found both a novel candidate biomarker and a candidate regulator. PIK3IP1, a lead signal in rapid decline and progression to KF, encodes a negative regulator of PI3K signaling^46^. Given the established role of PI3K/AKT signaling in podocyte insulin responsiveness and glucose uptake^47^, PIK3IP1 is a plausible but previously unexamined regulator of PI3K activity in DKD. Furthermore, in CKD, it was both overexpressed in connecting tubule cells (P=2.2×10^-13^) and underexpressed in endothelial cells of the glomerular capillary tuft (P=5.7×10^-7^). In the pQTL networks, TRIAP1 (TP53-regulated inhibitor of apoptosis 1), associated with progression to KF, was predicted as an upstream regulator of PPIB (Cyclophilin B) and MECR. Mouse models indicate that Triap1 protects against high glucose-induced podocyte apoptosis^49^, suggesting a protective response to kidney damage. PPIB plays a role in inflammation and fibrosis by modulating the response of tubular epithelial cells^48^, and MECR is involved in the mitochondrial fatty acid synthase pathway, regulated by ATP in hepatocytes, and associated with insulin resistance^51^. Together, these suggest TRIAP1 as a novel, central regulator of kidney health and PIK3IP1as a novel candidate biomarker.

Uromodulin (UMOD), the most abundant protein in urine, is produced in the thick ascending limb of Henle’s loop^48^. Consistent with previous reports^49^, lower serum UMOD was independently associated with rapid decline of kidney function, but not with the earlier albuminuria-based outcomes. Given its role in CKD^50^, UMOD may capture a distinct, albuminuria-independent component of rapid decline. In contrast, genetically predicted UMOD levels were associated with lower kidney function in MR. This may reflect differences between life-long genetically determined protein levels and circulating protein concentrations prior to kidney function decline.

In general, the integration of proteomic and genetic data provided orthogonal support of the biological specificity of measured protein measurements^17^. Whereas the longitudinal analyses identified a broad set, MR prioritized five proteins (UMOD, SORT1, CLSPN, MAST4 and EGF), whose genetically predicted levels were associated with eGFR in diabetes. The limited overlap between these likely reflects the different biological questions addressed by these approaches, the lack of robust genetic instruments for some proteins of interest and limited statistical power and availability of GWAS on eGFR and other DKD outcomes in diabetes. Longitudinal observational associations may capture early molecular perturbations or compensatory mechanisms, whereas MR identifies proteins whose genetically determined levels are linked to kidney function. Among the genetically prioritized proteins, the two kidney-tissue enriched proteins, UMOD and EGF, have established roles in kidney biology, and higher genetically predicted EGF levels were associated with higher eGFR in diabetes consistent with experimental studies on progressive renal injury and fibrosis^51^.

To our knowledge, the current study represents the largest, longitudinal proteomics study on diabetes outcomes with 5,420 proteins measured in 830 participants with type 1 diabetes and further validated in 249 independent individuals with either type 1 or type 2 diabetes; and one of the largest diabetes cohorts profiled at this scale. Further strengths include the dense clinical characterization and the long follow-up time. Our study has some limitations. The Olink platform provides relative protein quantification without transferrable thresholds for abnormal protein levels. For translation, targeted, quantified assays are needed. Although we detected *cis-*pQTLs for 60% of the proteins, without genetic instruments for many proteins, and with limited statistical power in the GWAS studies for eGFR in diabetes, most associations remain observational. Finally, both the discovery cohort and the main replication cohort consisted of Finnish individuals with type 1 diabetes. While we further investigated the lead protein associations in individuals with type 2 diabetes from Northern Ireland, these findings might not generalize to other ethnicities.

In conclusion, this large-scale proteomic analysis revealed that while there is a small number of shared, enriched pathways, distinct molecular processes characterized the successive stages of DKD progression in diabetes. Early disease was marked by inflammatory, kidney-associated and systematic stress-related signals (CD14, GFRA1, GDF15) detectable before clinically apparent kidney injury; albuminuria progression highlighted the kidney–liver axis (ASGR1, RBP4) linked to metabolic and inflammatory dysfunction; and advanced disease, namely rapid kidney function decline and KF, was characterized by proteins enriched for immune and TNF receptor signaling. Together with genetic support from pQTL and MR analyses, these findings extend our understanding of the underlying mechanisms of DKD and highlight opportunities for earlier risk stratification and therapeutic intervention.

## ACKNOWLEDGEMENTS

We are grateful to all individuals that took part in the FinnDiane Study and acknowledge the skilled technical assistance of Heli Krigsman, Hanna Olanne, Anna Sandelin, Anu Dufva and Kirsi Uljala (Folkhälsan Research Center, Finland), and all the physicians and nurses at each FinnDiane study centers taking part in the enrolment and clinical characterization of the participants (**Supplementary Table 15**). We would like to acknowledge support of SciLifeLab Affinity Proteomics at Uppsala University, Sweden and the National Genomic Infrastructure Uppsala (NGI), Sweden, supported by SciLifeLab, the Swedish Research Council and the Knut and Alice Wallenberg Foundation, for providing assistance in protein analyses. We want to acknowledge the ELIXIR Finland node hosted at CSC – IT Center for Science for ICT resources that enabled computational analysis. Furthermore, we wish to thank the Finnish Computing Competence Infrastructure (FCCI) for supporting this project with computational and data storage resources.

## AUTHOR CONTRIBUTIONS

NS and P-HG conceived the study and secured its funding. NS, P-HG, SM and ED designed the study. SM, ED, and XL performed the main data analysis. JK contributed to data processing and analysis. WJ and VN performed gene expression analyses and contributed to the writing. MK and KS contributed to the acquisition of the data and to its interpretation. ZA, HJ and AP performed replication analysis in type 2 diabetes, CH, LS, APM, AJM and GM contributed to the acquisition of the data and to its interpretation. EV, AS and VH contributed to outcome data creation. SM, ED, and NS wrote the first draft of the manuscript. All authors critically read, revised and approved the final version to be submitted and published manuscript.

## DATA AVAILABILITY

Individual-level data for the study participants are not publicly available because of the restrictions due to the study consent provided by the participant at the time of data collection.

Cross-sectional protein associations with cardio-kidney-metabolic traits are available in addition to **Supplementary Table 1** at https://finndiane.shinyapps.io/Protein_Association/.

pQTL associations will be made publicly available upon manuscript acceptance. Significant *cis*-pQTLs (FDR < 0.05) are in **Supplementary Table 12**.

Kidney bulk gene expression data are available in the public domain in Nephroseq (https://www.nephroseq.org) and the Gene Expression Omnibus (GEO) repository under the following: GSE104954 and GSE104948, and can be downloaded.

Kidney single-cell expression data are available from the Susztak Biolab (https://susztaklab.com/hk_genemap_kpmp/scRNA) and have been used (**Supplementary Table 7**) with written permission from Katalin Susztak (KS).

## CODE AVAILABILITY

No custom code or algorithm was developed for the analyses performed in this study, instead previously designed, publicly available, pipelines and code were used:

fgsea (version 1.30.0, https://bioconductor.org/packages/fgsea), ggraph (version 2.2.1, https://CRAN.R-project.org/package=ggraph), HPAanalyze (version 1.29.1, https://github.com/anhtr/HPAanalyze) logistf (version 1.26.1, https://CRAN.R-project.org/package=logistf), mgsigdbr (version 26.1.0, https://CRAN.R-project.org/package=msigdbr), OlinkAnalyze (version 4.1.2, https://CRAN.R-project.org/package=OlinkAnalyze), STRINGdb (version 2.16.4, https://bioconductor.org/packages/STRINGdb), survival (version 3.7-0, https://CRAN.R-project.org/package=survival), tidycmprsk (version 1.0.0, https://CRAN.R-project.org/package=tidycmprsk), UMAP (version 0.5.12, https://cran.r-project.org/package=umap), HDBscan (version 0.8.44, https://cran.r-project.org/package=dbscan), clusterProfiler (version 4.14.6, https://bioconductor.org/packages/release/bioc/html/clusterProfiler.html), ReactomePA (version 1.50.0, https://bioconductor.org/packages/release/bioc/html/ReactomePA.html), enrichplot (version 1.26.6, https://bioconductor.org/packages/release/bioc/html/enrichplot.html), org.Hs.eg.db R package (version 3.20.0, https://bioconductor.org/packages/release/data/annotation/html/org.Hs.eg.db.html), limma (version 3.62.2 (and version 3.58 for the differential analysis in kidney transcriptomic data in Supplementary Table 10), https://bioconductor.org/packages/release/bioc/html/limma.html), shiny (version 1.14.0, https://cran.r-project.org/package=shiny), MatrixeQTL (version 2.3, https://doi.org/10.32614/CRAN.package.MatrixEQTL). TwoSampleMR (Version: 0.7.8, https://mrcieu.github.io/TwoSampleMR/)

## METHODS

### Study cohort and outcomes

This study included 830 adults with type 1 diabetes from the FinnDiane Study. T1D was defined as T1D diagnosed by their attending physician, age of diabetes onset below 40 years of age, and requiring insulin in the first year after diagnosis. All participants underwent a thorough clinical examination at the study visit and provided blood and urine samples. Baseline albuminuria status was assigned based on agreement in two out of three measurements of the AER. We selected for proteomics analysis 449 participants with no kidney disease at the study baseline (normal AER and eGFR > 60 mL/min/1.73m^2^), 149 participants with moderate albuminuria, and 232 with severe albuminuria at baseline. Within each of these three sub-cohorts, we included individuals with confirmed stable albuminuria status for at least 5 years of follow-up and individuals with deteriorating kidney health during the follow-up, and matched these two groups based on sex, baseline age and diabetes duration, and the storage time and number of thaws of baseline serum samples (all p ≥ 0.05). Individuals with normal AER and moderate albuminuria were additionally matched for baseline eGFR (p > 0.05). Among those with normal AER at baseline, we further required preservation of eGFR (> 60 mL/min/1.73m2) at the end of follow-up, for inclusion in the stable group. Additionally, we created a fourth sub-cohort of 730 adults with available data on eGFR slopes (**Supplementary Fig. 1**). Slopes were calculated from the study visit forward based on serum creatinine measurements using the CKD-EPI 2021 formula^52^. We included those individuals with a first eGFR observation within three months of the study visit and least three eGFR observations in at least two years of follow-up. Slope calculation stopped at CKD stage 5 (defined as first eGFR<15mL/min/m^2^ and a confirmatory eGFR within 30 to 365 days), at KF (with a presumed final eGFR of 10mL/min/m^2^), or at the last available measurement, whichever came first. All slopes were calculated based on calendar month median eGFRs. All slopes were visually inspected and one slope was removed due to quality issues.

We investigated the following longitudinal outcomes:

- For those without kidney disease at the time of sampling (N=449), onset of albuminuria (N=165) over a follow-up time of 10.2 ± 5.4 years
- For those diagnosed with moderate albuminuria at the time of sampling (N=149), progression to severe albuminuria (N=60) over a follow-up time of 8.5 ± 5.0 years
- For those diagnosed with severe albuminuria at the time of sampling (N=232), progression to KF (N=104) over a follow-up time of 9.0 ± 5.2 years
- For all adults with eGFR slopes available (N=730), rapid decline of kidney function after baseline, defined as an annual eGFR loss of more than 3 mL/min/1.73m^2^ (N=145).

The study protocol was approved by the HUS Regional Committee on Medical Research Ethics (491/E5/2006, 238/13/03/00/2015, and HUS/3313/2018), and the study was performed in accordance with the Declaration of Helsinki. Written informed consent was obtained from each participant.

### Serum sample pre-processing and proteomics measurements

The median number of thaw cycles was 1 (IQR 1 - 2) with 20 % of samples that have never been thawed before. The maximum number of thaw cycles was 5. The samples were randomized on plates based on sex, age, baseline albuminuria status, kidney disease progression, storage time and thaw cycle number. Altogether 840 serum samples that were stored in –20° Celsius were sent for proteomic analysis at SciLifeLab, Uppsala, Sweden. Serum protein levels were quantified using the Olink Explore HT, which combines proximity extension assays with next-generation sequencing and measures 5,420 proteins. The protein expression data was intensity normalized by the SciLifeLab. Of the samples, three failed the laboratory quality control and were removed. A further seven samples were removed as they were considered outliers with either their median or IQR protein concentrations exceeding 3.5 standard deviations of the overall median or IQR protein concentration (using the OlinkAnalyze R package). Three of those were also considered outliers in a Principal Component Analysis (PCA) plot (**Supplementary Fig. 3**). Due to careful sample randomization for the plates, the protein expression data was not batch adjusted.

### Multiple testing corrections

To account for multiple testing in the proteome-wide analyses, we used Bonferroni correction applied to the independent numbers of tests. The effective number of independent tests was calculated using PCA, which showed that the first 637 principal components cover 99% of the variance in the protein abundance data. The P value threshold for multiple comparisons was therefore adjusted by dividing 0.05 by 637 resulting in a proteome-wide significant threshold of P=7.85×10^-5^.

### Assessment of protein stability during storage

The serum for the 830 samples was stored in –20° Celsius for a median of 20.0 years (IQR 16.2 - 22.3 years). The maximum storage time was 25.2 years. Protein stability was evaluated in a subset of the participants (n=20). For each participant, two aliquots collected from the same study visit were stored in parallel at either –20 °C or at –80 °C. For each protein, protein abundance measured in samples stored at −20 °C was compared with that in paired samples stored at −80 °C with Pearson and Spearman correlation coefficients used to assess the linear relationship and rank order correlation, respectively.

### Proteomic clustering

All 5,420 proteins were assigned to different clusters based on the NPX levels. UMAP (version 0.5.12) algorithm was used for dimension reduction, followed by Hierarchical Density-Based Spatial Clustering of Applications with Noise (min_cluster_size=120, min_samples=15; HDBSCAN version 0.8.44) clustering based on UMAP coordinates. The silhouette score was used to determine clustering performance. Reactome pathway enrichment analysis and visualization were performed with the clusterProfiler (version 4.14.6), ReactomePA (version 1.50.0), and enrichplot (version 1.26.6) R packages. All proteins were mapped to Entrez Gene identifiers with org.Hs.eg.db R package (version 3.20.0) and were used as background set. Significant enriched pathways were defined as pathways with Benjamini–Hochberg-adjusted P < 0.05. Up to five most significant pathways were visualized for each cluster.

### Cross-sectional outcomes

Associations between protein abundance (NPX levels: dependent variable) and baseline clinic characteristics (independent variables) were assessed using multiple variable linear regression models adjusted for sex, age, storage time and thaw number. Clinical characteristics included hemoglobin A1c (hba1c), duration of type 1 diabetes (T1D duration), body mass index (BMI), waist-height ratio (WHtR), high-sensitivity C-reactive protein (Hs-CRP), total cholesterol, low-density lipoprotein cholesterol (LDL Chol), blood triglyceride level (Triglycerides), current smoking status (smoking), and systolic blood pressure (SBP). We repeated the association analysis with baseline eGFR as an additional covariate.

Given some evidence of quasi-complete separation (when using standard logistic regression models) and the smaller sample size, the associations between baseline NPX and severe albuminuria (normal AER or moderate albuminuria vs. severe albuminuria) and additionally with baseline CKD stage 3 or 4 (eGFR <60mL/min/1.73m^2^) were assessed with Firth’s bias-reduced logistic regression models (penalization of the log-likelihood by Jeffreys prior) using the logistf R package^53^. The minimal models were adjusted for sex, storage time and thaw numbers whereas the adjusted models additionally also for either eGFR (albuminuria analysis) or severe albuminuria (CKD analysis).

### Longitudinal kidney outcomes

Albuminuria onset, its progression, and progression to KF were assessed using a competing risk framework. For all those three outcomes, death from all causes was treated as a competing event. Participants who experienced neither event were censored at the date of last follow-up. Associations were evaluated using the Fine-Gray sub-distribution hazard model (sHR) (survival R package^54^). The proportional hazard assumption was tested for all models and if a specific protein violated the assumption, the follow-up time was reduced to 10 years and if necessary, further reduced to 5 years. The protein associations with rapid decline of kidney function were assessed with Firth’s bias-reduced logistic regression. Minimal models in all longitudinal analyses were adjusted for storage time, thaw numbers and sex. For the progression to KF, minimal models were additionally adjusted for eGFR, and the minimal models for rapid kidney function decline were additionally adjusted for eGFR and severe albuminuria. The fully adjusted models were adjusted for storage time, thaw number, sex, age, age of diabetes onset, eGFR, HbA1c, triglycerides, systolic blood pressure, waist-height ratio, hs-CRP and current smoking status. Fully adjusted models for rapid decline of kidney function were additionally adjusted for severe albuminuria. We report the results of fully adjusted models in the main text. The tidycmprsk R package was used to create cumulative incidence curves.

### Functional enrichment analysis

Gene set enrichment analysis (fgsea package^55^) using the proteins’ gene names was performed to infer altered pathways. For each cross-sectional and longitudinal regression analysis, proteins were sorted according to their t-statistics (β coefficient divided by standard error). In the cross-sectional analyses, the t-statistics were based on the results of the adjusted analyses and for the longitudinal analyses, they were based on minimal models. Reactome^56^ gene sets for humans (N=1839) were retrieved from MSigDB (release 2026.1.Hs) using the msigdbr package (version 2026.1.0)^57^. The minimum number of overlaps between the tested set and a Reactome gene set was set to 15, the maximum to 500 and the number of permutations to 100,000. Pathways with an FDR below 0.05 were collapsed into independent ones.

All analyses were performed in R using version 4.4.2.

### Tissue expression and correlation with clinical variables

Using the HPAanalyze R package^58^, we downloaded tissue expression data from the Human Protein Atlas (Version 25, accessed 2026-08-09)^59,60^ and removed entries at the level *Uncertain*. For each tissue and protein, we reported the expression level of the most highly expressed cell type in that tissue and for that protein (levels are ordered from lowest to highest: not detected – low – medium – high). From the 59 replicated top proteins, 41 had detectable protein expression. We used hierarchical clustering for both the tissues and the proteins in the heatmap (**Extended Data Fig. 4**).

Additionally, we created heatmaps for the Spearman correlation between baseline clinical markers and the 59 replicated, top proteins. Again, we used hierarchical clustering for both the proteins and the clinical markers (**Extended Data Fig. 3**).

### Protein-protein interaction network analysis

Protein-protein interaction (PPI) data were retrieved from the STRING database (Version 12, accessed 2026-08-09) restricted to *Homo sapiens* using the STRINGdb R package^61^. Interactions were queried for the 59 replicated proteins using a minimum required interaction confidence score of 400 (medium confidence) in this exploratory analysis. Prior to network construction, the statistical significance of protein-protein interaction enrichment was assessed using STRING’s PPI enrichment test. A network was considered to reflect greater connectivity than expected by chance at p < 0.05. Network visualization was performed using ggraph. Node size was scaled to reflect degree centrality, and edge width was scaled to the STRING interaction confidence score (**Extended Data Fig. 5**).

### Replication analyses in type 1 diabetes

Replication analyses were performed in an independent validation cohort consisting of 122 individuals with type 1 diabetes from the FinnDiane study. Proteins were quantified using the same Olink Explore HT platform as in the discovery cohort, but in plasma instead of serum and in a different laboratory at the Austrian Institute of Technology (Austria). The plasma samples had been stored at –80 degrees Celsius and were never thawed previously. The proteomics data was processed in the same way as the data in the discovery cohort.

Replication analyses were performed using minimally adjusted (sex and storage time cut into tertiles) Firth’s bias-reduced logistic regression models due to the limited size of the replication cohort. As longitudinal follow-up data were only available for a subset of participants (64 of 122), proteins associated with the longitudinal outcomes in the discovery cohort (albuminuria onset, albuminuria progression, rapid KF), were evaluated against analogous baseline kidney phenotypes in the replication cohort. Specifically, proteins associated with progression to albuminuria onset (n=5), were evaluated against prevalent albuminuria at baseline (moderate or severe vs no albuminuria). Proteins associated with albuminuria progression (n=21), were evaluated against severe albuminuria, whereas proteins associated with rapid kidney function decline (n=34), were evaluated against baseline CKD stage 3 or 4 (eGFR < 60mL/min/1.73m^2^) due to a lack of longitudinal slope data. Among the 64 individuals with longitudinal follow-up data available, a composite endpoint of KF or ≥40% eGFR decline occurred in 12 individuals during a median follow-up time of 2.9 years (233 person-years). The proteins associated with progression to KF (n=98) were evaluated against this composite endpoint using minimally adjusted (sex and storage time) Fine Gray analyses, with death included as a competing event. The proportional hazard assumption was assessed and satisfied for all replication models.

For each outcome, the Bonferroni-corrected P value threshold was calculated by dividing 0.05 by the number of proteins evaluated in the corresponding replication analysis, i.e. P<0.01 for moderate or severe albuminuria; P<0.0024 for severe albuminuria; P<5.1×10^-4^ for albuminuria progression to KF; and P<0.0015 for rapid decline.

### Kidney tissue transcriptomic analysis

Differential kidney gene expression in DKD was analyzed using microdissected glomerular (n=31 and 12) and tubulointerstitial compartment data (n=30 and 17) from healthy living donors and patients with DKD, respectively, of the European cDNA biobank^14–16^ for 15 proteins (**Supplementary Table 10**). The limma method (Linear Models for Microarray Data) was used to identify differentially regulated genes.

### Evidence in kidney single-cell RNA sequencing data

To find further evidence for kidney expression, we queried single cell RNA transcriptomics data in in healthy controls and individuals with CKD from the Human Kidney Single Cell Transcriptome dataset^13^ (*Susztak & KPMP merged*, https://susztaklab.com/hk_genemap_kpmp/scRNA, **Supplementary Table 7**) for the 59 replicated proteins. Log and normalized average expressions are reported across all samples, control samples, and CKD samples. Wilcoxon rank-sum test was used to assess if each gene expression is differentially expressed for each cell type, and differentially expressed in CKD samples as compared to control samples. P-values were corrected for multiple testing using the Benjamini-Hochberg FDR method (adjusted for testing 34,733 genes).

### External validation in type 2 diabetes cohort

We evaluated the 59 replicated proteins in 127 individuals with type 2 diabetes and eGFR<60mL/min/1.73m^2^ from the NICOLA cohort (**Supplementary Table 9**). Protein measurements were generated simultaneously with the 122 samples from the type 1 diabetes replication cohort, and protein measurements were generated using the same Olink platform and analytical workflow. Associations between protein levels and kidney function were assessed using two complementary outcomes: 1) continuous eGFR and 2) advance DKD, defined as eGFR <45 mL/min/1.73m^2^ (CKD stage 3b or worse). Associations with eGFR were analysed using linear regression, whereas associations with advanced DKD were evaluated using logistic regression. All models were adjusted for age, sex and Olink plate. Multiple testing was performed using the Benjamini-Hochberg FDR method.

### pQTL analysis

To identify the genetic determinants for proteins on the Olink Explore HT panel, we performed a pQTL analysis with MatrixEQTL software (v 2.3). We analyzed both local (cis) and distant (trans) effects, using a threshold of 1 Mb for detection of cis–pQTLs. We used imputed genome-wide genotyping data genotyped in four batches at the University of Virginia using HumanCoreExome Bead arrays (versions 12-1.0, 12-1.1 and 24-1.0, Illumina, San Diego,CA) and imputed using the population-specific Finnish reference panel (SISu version 3). and included only variants imputed with high imputation quality (INFO>0.80) and MAF>0.01.

A pQTL-supported protein regulatory network was then constructed for the 130 proteome-wide associated proteins. Trans-pQTL variants was restricted to those located on a different chromosome or more than 5Mb from target gene TSS. For each protein, downstream targets were defined as proteins associated in *trans* with SNPs with leading *cis*-pQTL variant (FDR<0.1, MAF>=0.05). Upstream regulators were defined as proteins lead *cis*-pQTL variant were associated in *trans* with other proteins (FDR<0.1, MAF>=0.05). Network is visualized with Cytoscape (version: 3.10.2).

### MR analysis

We performed MR analyses to investigate potential causal relationships between circulating protein levels and diabetic kidney disease. Proteins with at least one significant pQTL identified in our pQTL analysis (cis-pQTLs at FDR<0.05 and trans-pQTLs at FDR<0.05) and a MAF≥ 0.05 were considered eligible instrumental variables.

Two-sample MR (TwoSampleMR R package) was performed using both cis- and trans-pQTLs as instrumental variables. When only a single genetic instrument was available, causal effects were estimated using the Wald ratio method; otherwise, inverse-variance weighted (IVW) MR was applied.

As outcomes, we used summary statistics from GWAS on eGFR in individuals with diabetes, including: all individuals with diabetes, (n=36,169) and in individuals without diabetic kidney disease (defined as severe albuminuria or worse, n=24,220), as described by Cole et al. (2025)^19^. Diabetes was defined as type 1 diabetes or type 2 diabetes with any diabetes duration (“Diabetes Loose” analysis).

## EXTENDED DATA FIGURES

**Extended Data Figure 1.**
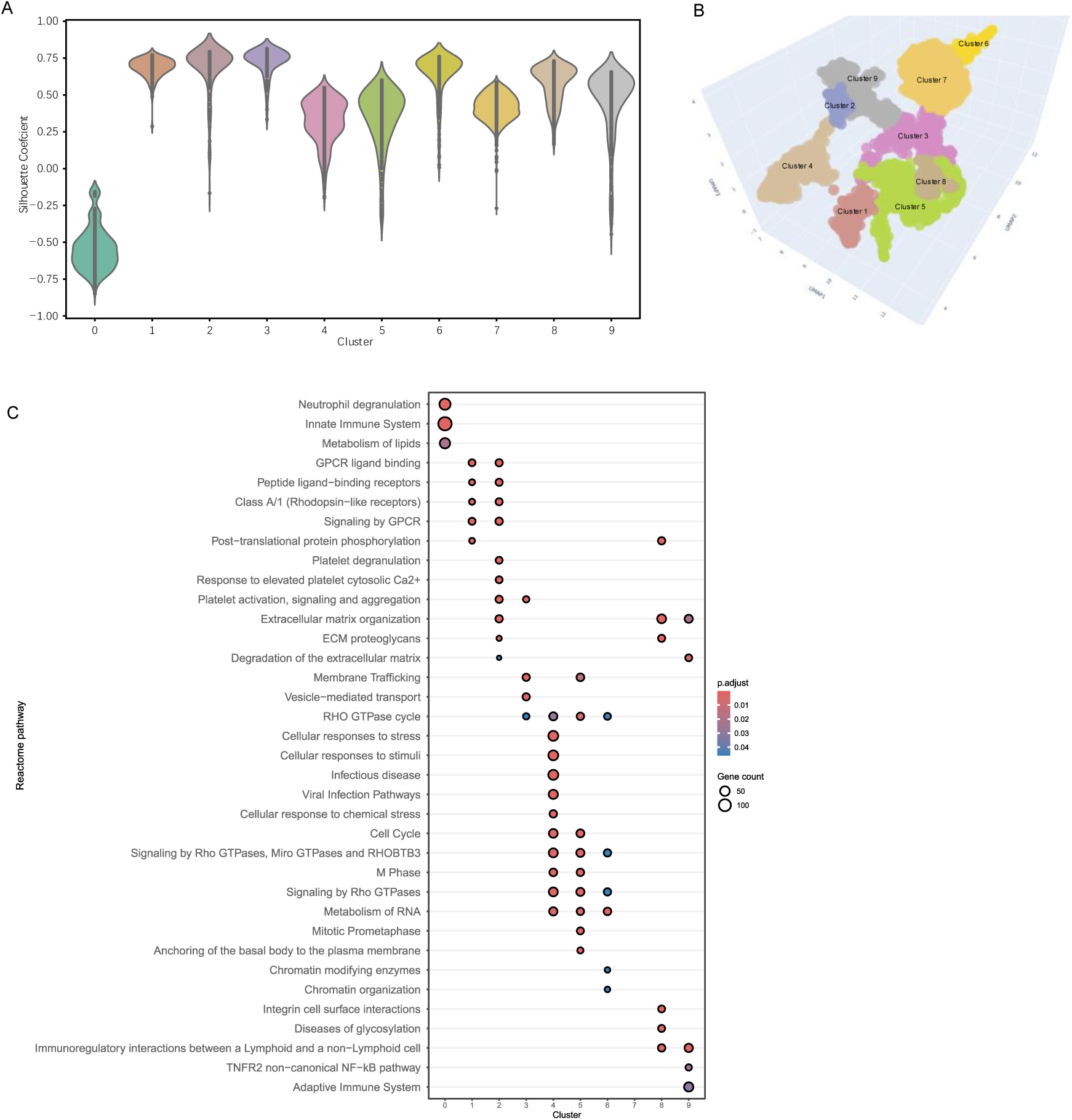
**(A)** Silhouette score distributions of all protein clusters; **(B)** 3D UMAP of protein clusters; **(C)** Reactome pathway enrichment of proteins in different clusters. **Cluster 1**, with 185 proteins, was enriched in G protein-coupled receptor (GPCR)-related pathways, including GPCR ligand binding, peptide ligand−binding receptors signaling by GPCRs and Class A/1 rhodopsin-like receptors, GPCR signaling, and post-translation protein phosphorylation. 145 proteins in **cluster 2** were enriched in GPCR-related pathways and platelet-associated processes, including platelet degranulation, response to elevated platelet cytosolic Ca2+, platelet activation, signaling and aggregation, together with extracellular matrix organization. **Cluster 3,** with 142 proteins, was enriched in membrane trafficking, vesicle-mediated transport, and RHO GTPase cycle. **Cluster 4**, with 542 proteins, was mainly associated with a broader functional pathway, including cellular stress and stimulus response, infection-related pathways, cell-cycle processes, RHO GTPase signaling, and RNA metabolism. **Cluster 5**, with 362 proteins, was enriched in cell-cycle and RHO GTPase-related pathways. 314 proteins in **Cluster 6** were enriched in RHO GTPase signaling, RNA metabolism, and chromatin modifying enzymes. **cluster 8,** with 584 proteins, was enriched in immunoregulatory interactions between a lymphoid and a non−Lymphoid cell, glycosylation disease, and integrin cell surface interactions. **Cluster 9** was enriched in immunoregulatory interactions between a lymphoid and a non−Lymphoid cell, non-canonical NF-KB pathway, and adaptive immune system. **Cluster 7**, with 1,467 proteins, does not exhibit any significant pathway enrichment.

**Extended Data Figure 2.**
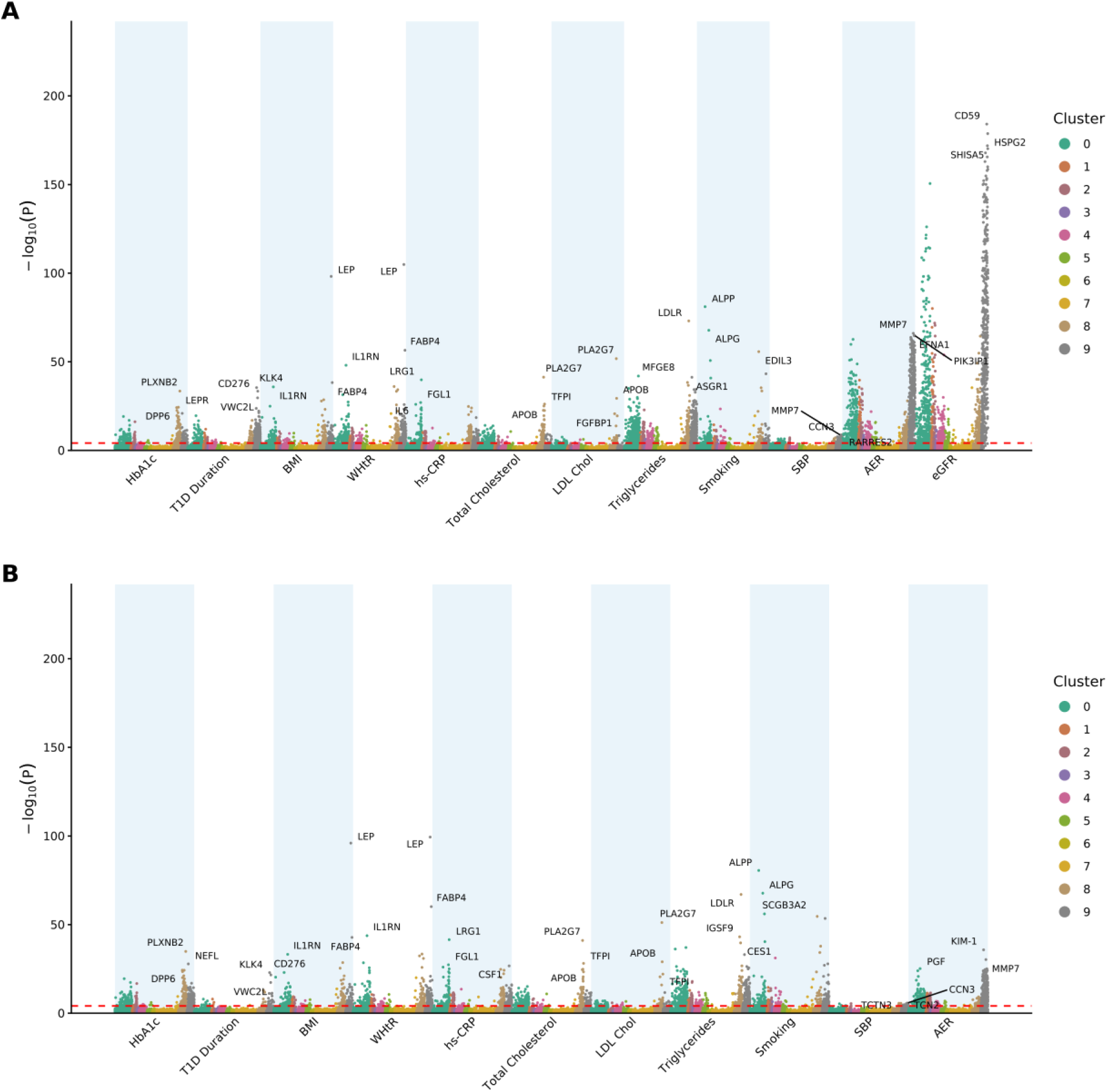
Cross-sectional associations between circulating proteins and clinical traits at baseline (A) and after additionally adjusting for baseline estimated glomerular filtration rate (eGFR) (B). Associations between circulating proteins and clinical variables at baseline are shown as -log10(*p*)-values. Analyses were adjusted for age, sex, thaw number, storage time and eGFR. Proteins are sorted on the x-axis according to the unsupervised clustering of the proteins. The dashed line represents the Bonferroni-corrected threshold for number of independent proteins (*p* < 7.84 × 10^-5^). For each clinical trait, the three most significant protein associations are labeled. HbA1c: hemoglobin A1c, T1D Duration: duration of type 1 diabetes, BMI: body mass index, WHtR: waist-height-ratio, hs-CRP: high-sensitivity C-reactive protein, LDL cholesterol: low-density lipoprotein cholesterol, SBP: systolic blood pressure, AER: albumin excretion rate.

**Extended Data Figure 3.**
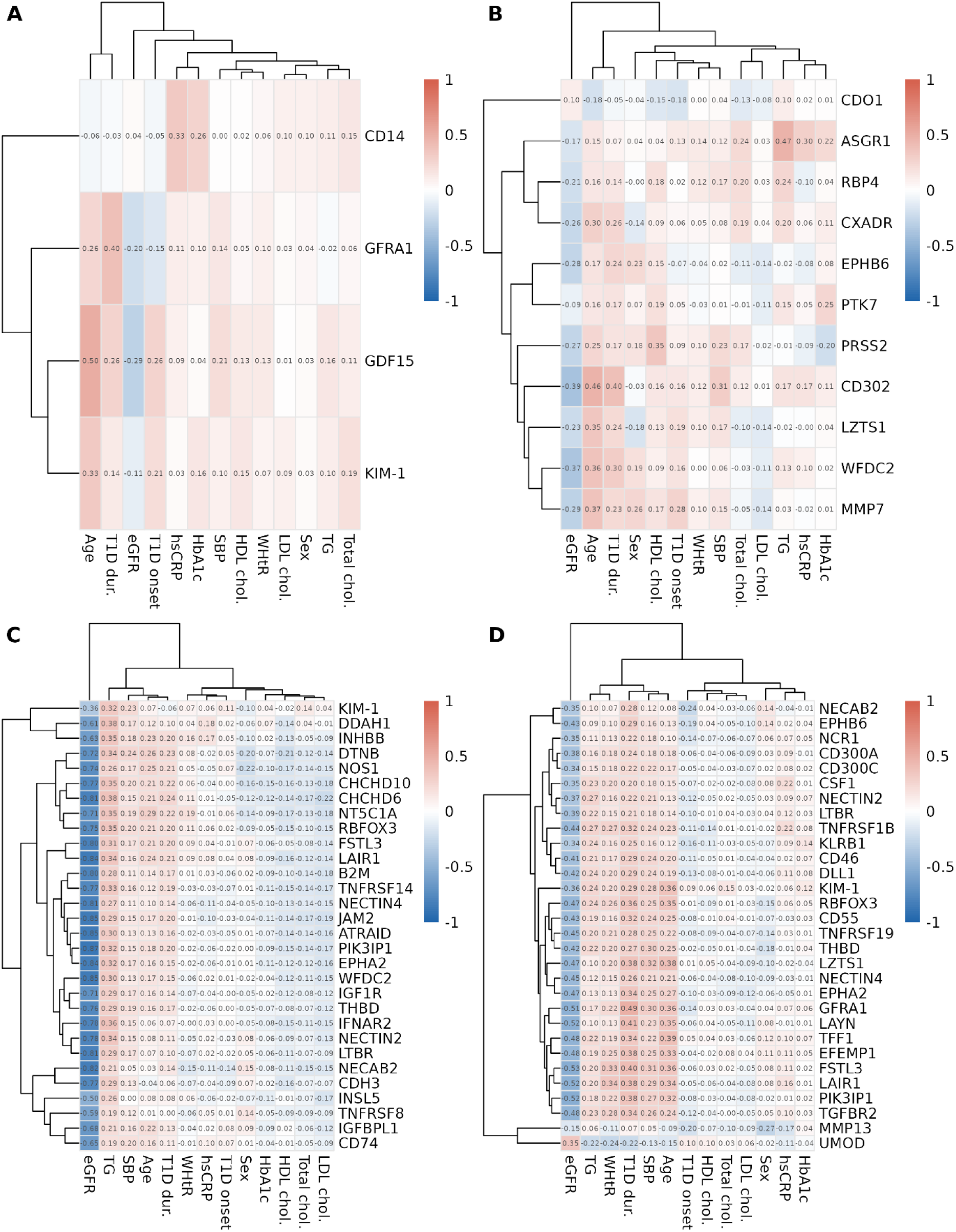
Heatmaps comparing baseline clinical markers to baseline protein NPX values for the 59 replicated proteins using Spearman correlation. (A) Albuminuria onset, (B) albuminuria progression, (C) progression to kidney failure and (D) rapid decline of kidney function. Total chol.: total cholesterol, HbA1c: glycated hemoglobin A1c, hsCRP: high-sensitivity C-reactive protein, LDL chol.: low density lipoprotein cholesterol, HDL chol.: high density lipoprotein cholesterol, T1D onset: age of type 1 diabetes onset, SBP: systolic blood pressure, T1D dur.: duration of type 1 diabetes, WHtR: waist-height ratio, TG: triglycerides, eGFR: estimated glomerular filtration rate

**Extended Data Figure 4.**
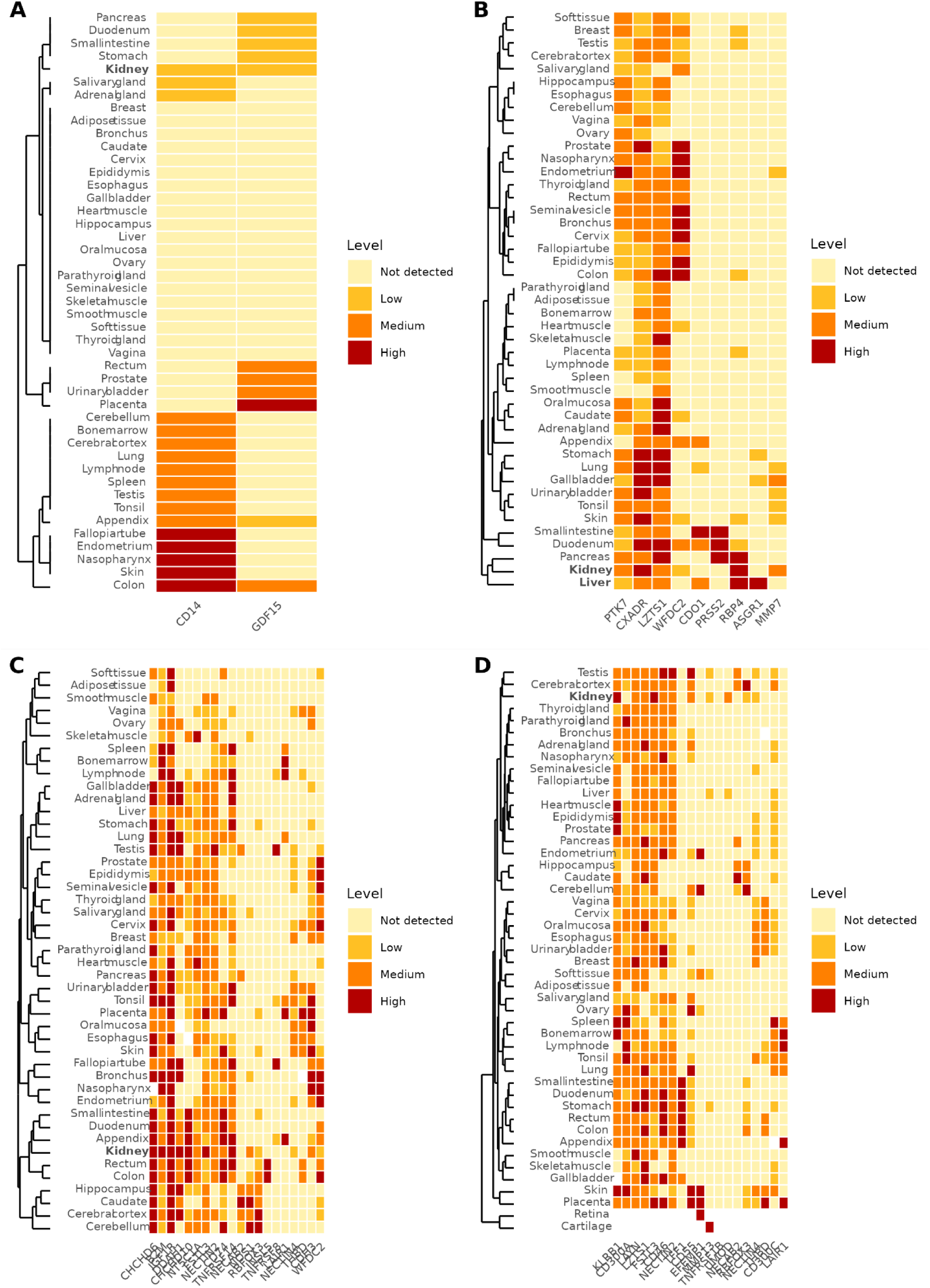
Protein expressions of top hits from the Human Protein Atlas (version 25) for onset (A), progression (B), kidney failure (C) and rapid decline (D) excluding those with level *Uncertain*.

**Extended Data Figure 5.**
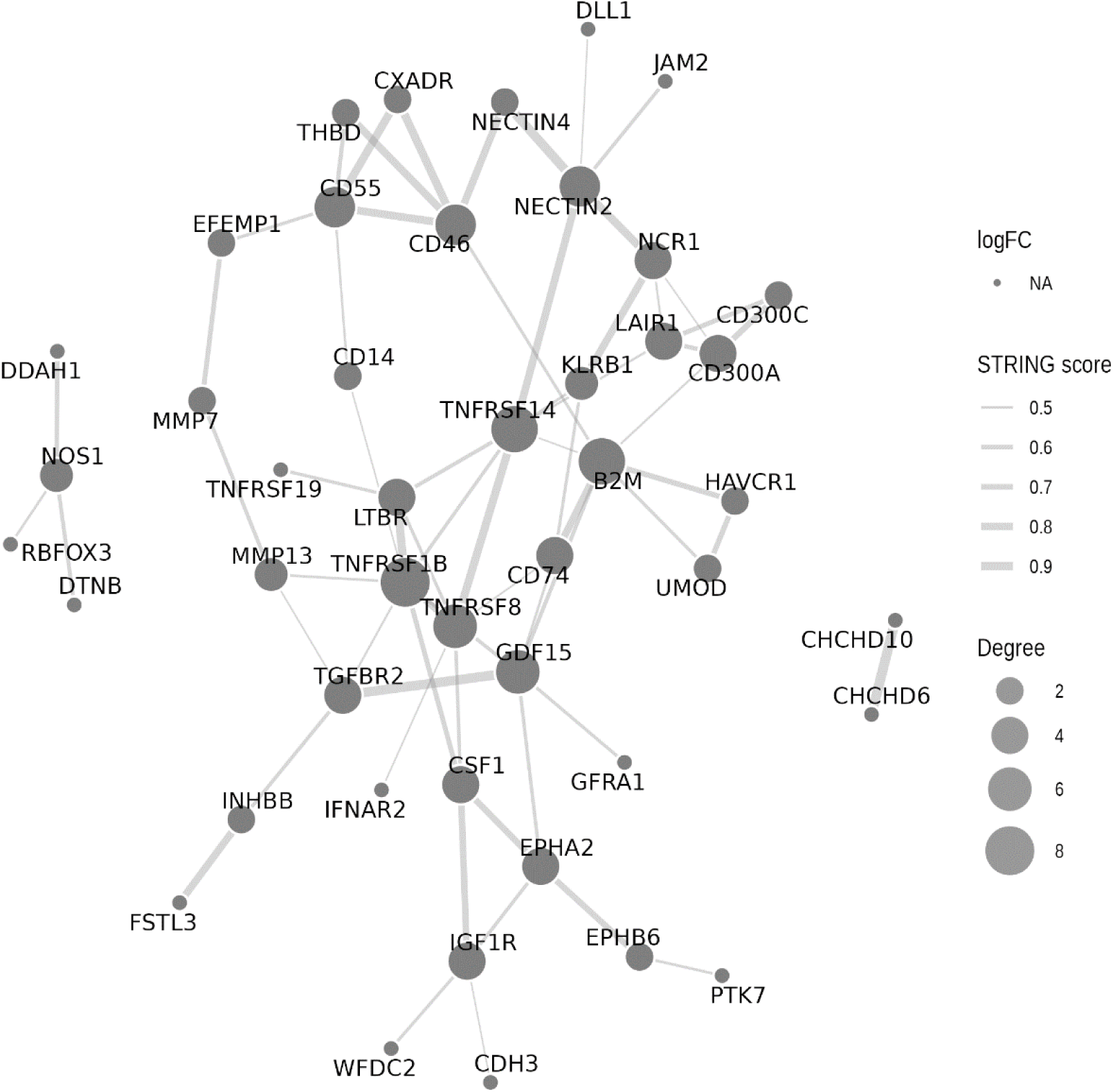
Protein-protein interaction network with STRINGDb (Version 12) for the 59 proteins that were associated with any of the four longitudinal DKD outcomes and replicated.

**Extended Data Figure 6.**
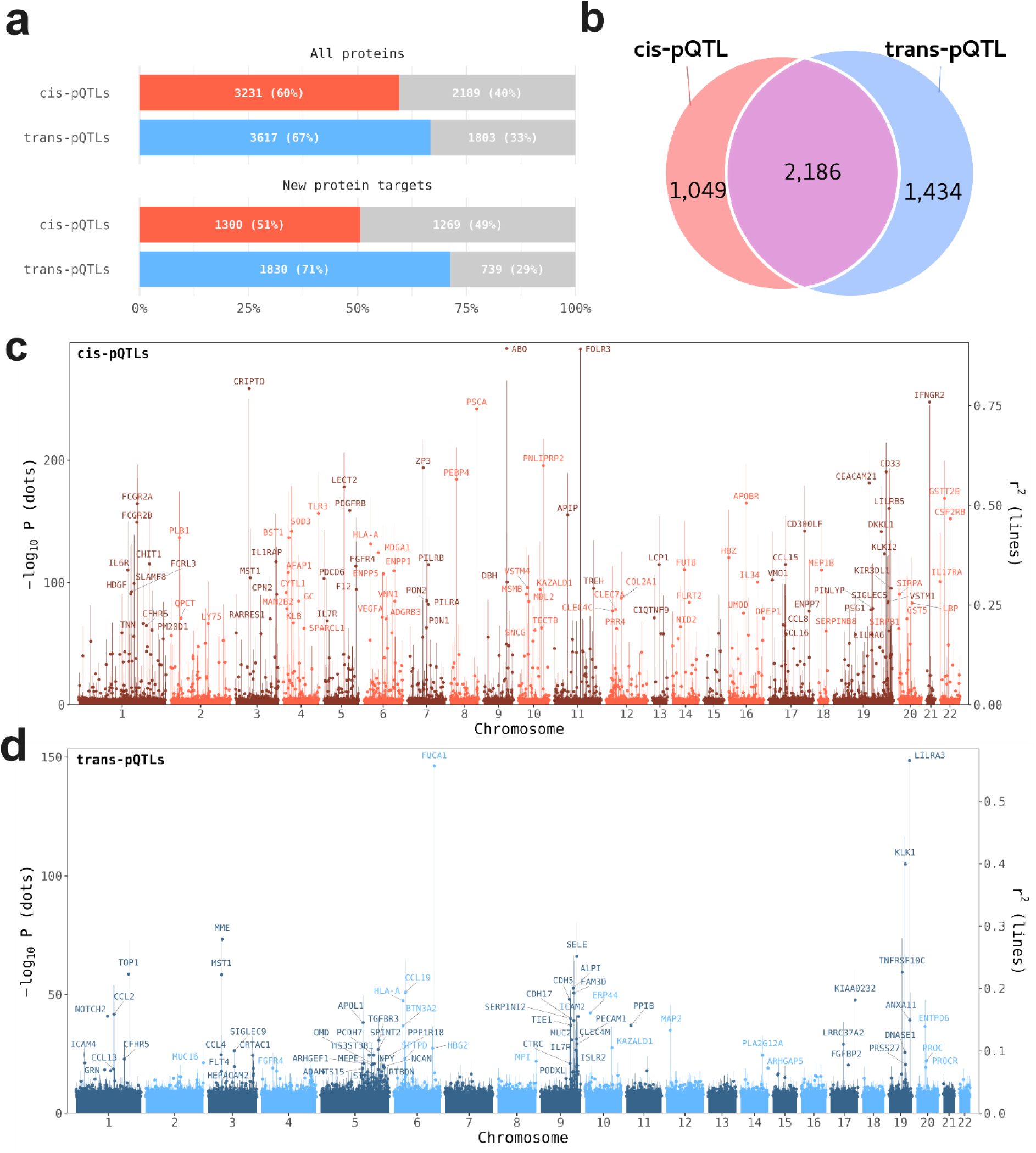
Landscape of cis- and trans-pQTLs identified across circulating proteins on Olink Explore HT. A, proportion of proteins with at least one significant cis-pQTL or trans-pQTL among all proteins analyzed and among newly included protein targets on ExploreHT. Numbers within bars indicate the number and percentage of proteins with (colored) and without (grey) pQTLs. B, overlap between proteins harboring cis- and trans-pQTLs. C, D, Genomic distribution of significant cis-pQTLs (c) and trans-pQTLs (d) Each point represents the lead variant per protein, plotted according to chromosomal position and associations significance (-log10 P). Vertical lines indicate the proportion of variance in protein abundance explained by each variant (r^2^, right axis). Selected variants labelled by the measured protein.

**Extended Data Figure 7.**
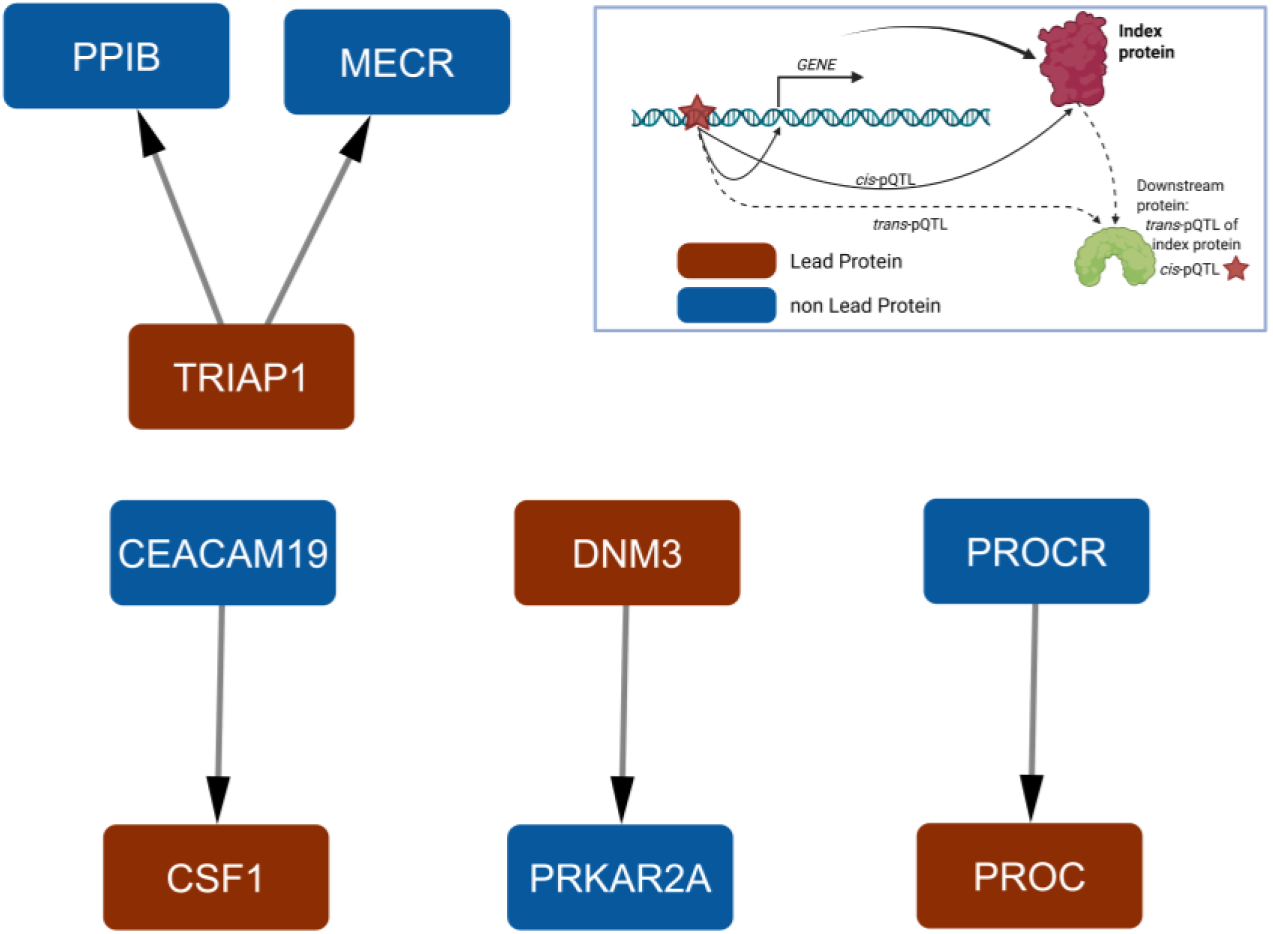
Putative regulation of DKD-associated proteins inferred from pQTL associations. Putative regulation was identified based on shared significant cis- and trans-pQTL variants (FDR<0.1, MAF≥0.05, trans-pQTL variants located on a different chromosome or outside 5Mb of the gene TSS). Arrows indicate that the *cis*-pQTL of the upstream protein was also associated in trans with the downstream protein. Red nodes represent lead proteins, and blue nodes represent non-lead proteins. Schematic diagram of the putative regulatory mechanism is shown in the top-right panel.

