## Supplementary Figures for "Comprehensive proteomics reveals stage-specific signatures in blood that precede diabetic kidney disease onset and progression"

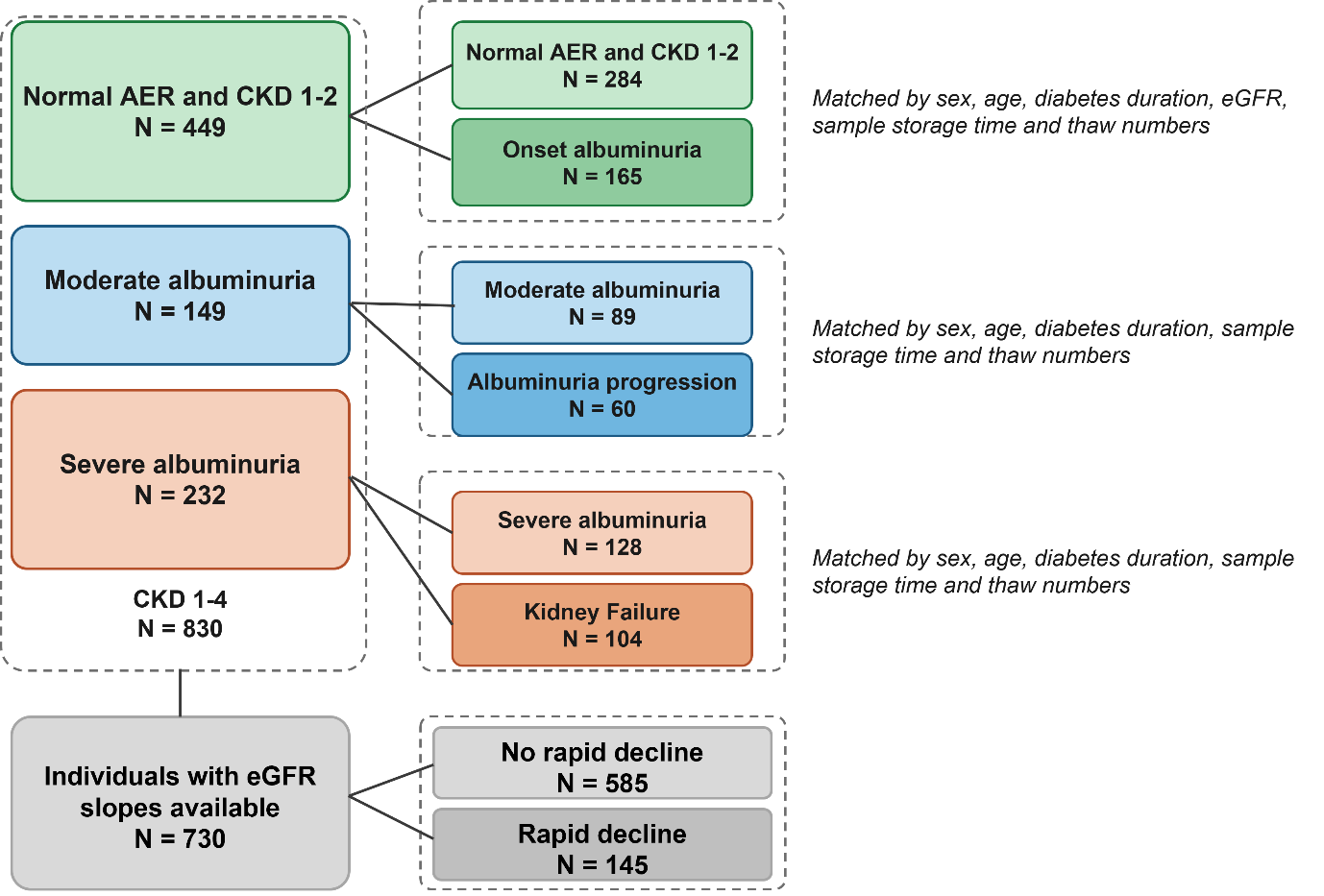

**Supplementary Figure 1.** Study Design for 830 adults with type 1 diabetes and how they are assigned to the four longitudinal outcomes. AER: Albumin excretion rate, CKD: chronic kidney disease, eGFR: estimated glomerular filtration rate

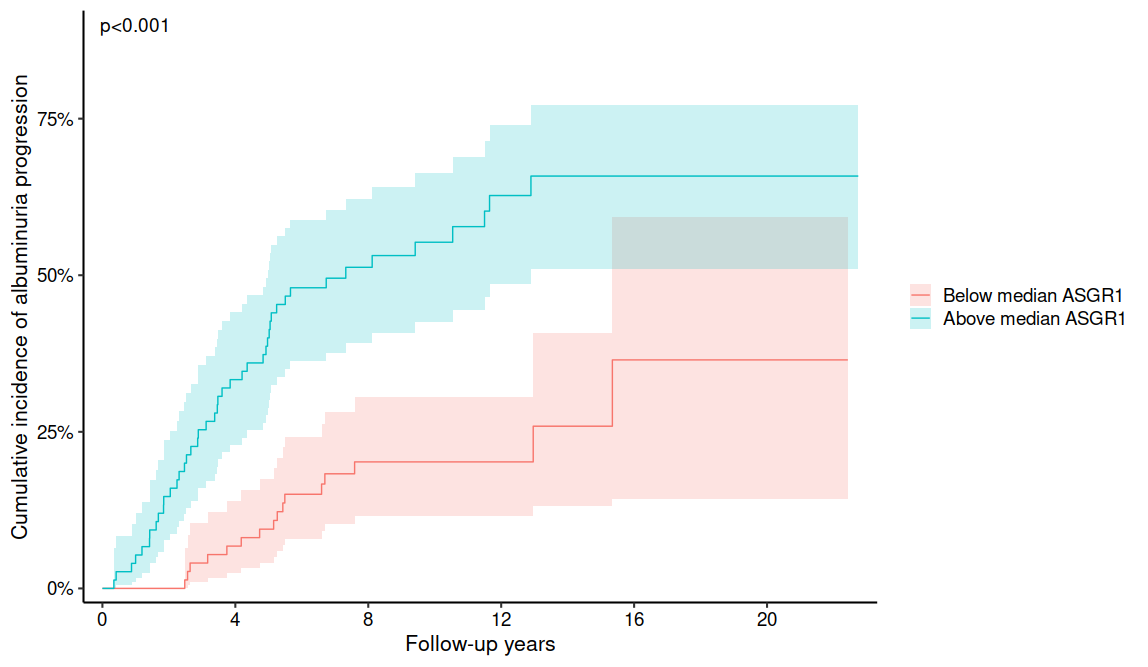

**Supplementary Figure 2**. Cumulative incidence plot for progression in albuminuria split between those with ASGR1 abundance above and below the cohort median

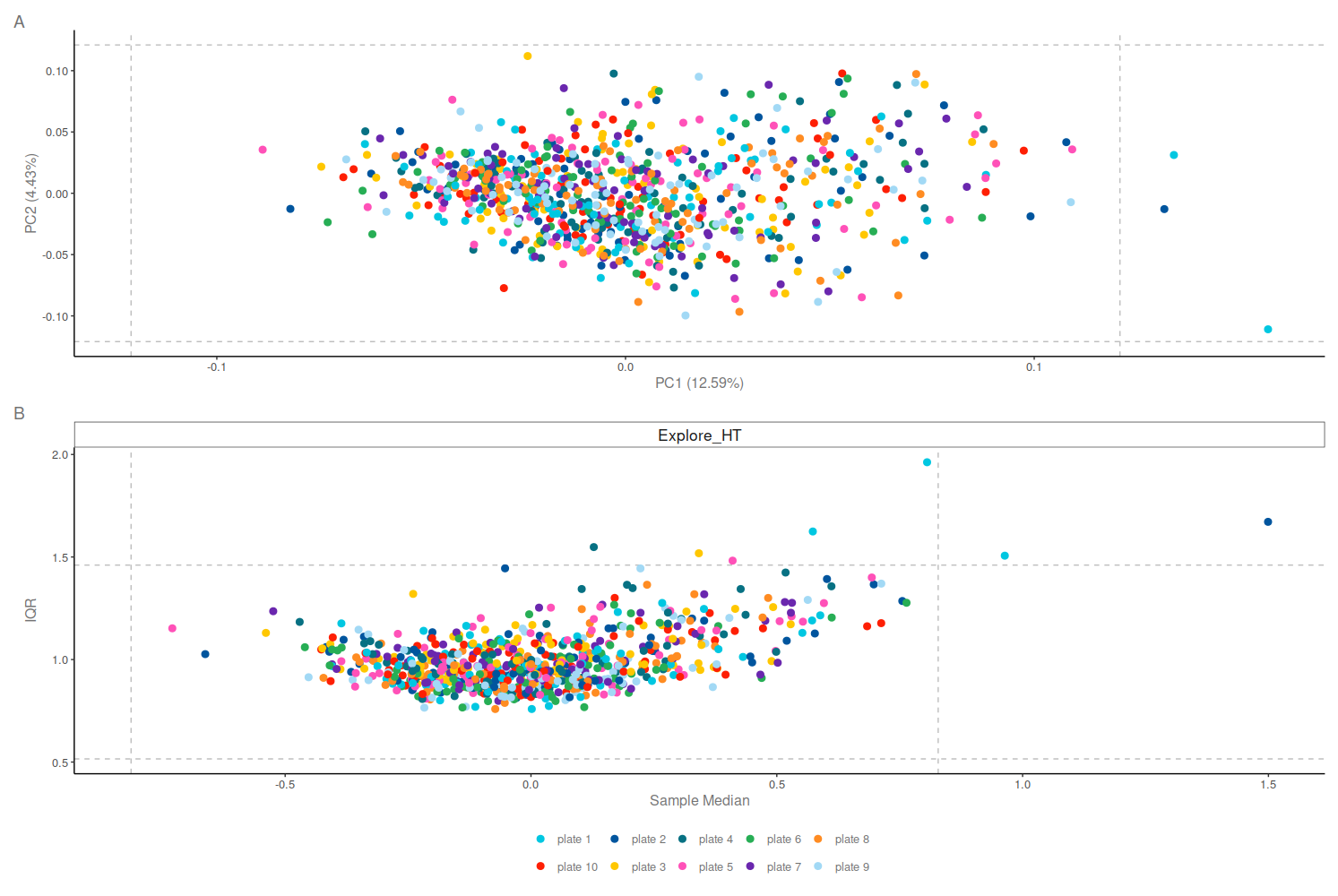

**Supplementary Figure 3**. Outlier detection during quality control: (A) By PCA plot and (B) by plotting the sample median vs. interquartile range (IQR). Outliers were defined as being more than 3.5 standard deviations away (dotted lines).
